# Hematopoietic mosaic chromosomal alterations and risk for infection among 767,891 individuals without blood cancer

**DOI:** 10.1101/2020.11.12.20230821

**Authors:** Seyedeh M. Zekavat, Shu-Hong Lin, Alexander G. Bick, Aoxing Liu, Kaavya Paruchuri, Md Mesbah Uddin, Yixuan Ye, Zhaolong Yu, Xiaoxi Liu, Yoichiro Kamatani, James P. Pirruccello, Akhil Pampana, Po-Ru Loh, Puja Kohli, Steven A. McCarroll, Benjamin Neale, Eric A. Engels, Derek W. Brown, Jordan W. Smoller, Robert Green, Elizabeth W. Karlson, Matthew Lebo, Patrick T. Ellinor, Scott T. Weiss, Mark J. Daly, The Biobank Japan Project, FinnGen Consortium, Chikashi Terao, Hongyu Zhao, Benjamin L. Ebert, COVID-19 Host Genetics Initiative, Andrea Ganna, Mitchell J. Machiela, Giulio Genovese, Pradeep Natarajan

## Abstract

Age is the dominant risk factor for infectious diseases, but the mechanisms linking the two are incompletely understood^1,2^. Age-related mosaic chromosomal alterations (mCAs) detected from blood-derived DNA genotyping, are structural somatic variants associated with aberrant leukocyte cell counts, hematological malignancy, and mortality^3-11^. Whether mCAs represent independent risk factors for infection is unknown. Here we use genome-wide genotyping of blood DNA to show that mCAs predispose to diverse infectious diseases. We analyzed mCAs from 767,891 individuals without hematological cancer at DNA acquisition across four countries. Expanded mCA (cell fraction >10%) prevalence approached 4% by 60 years of age and was associated with diverse incident infections, including sepsis, pneumonia, and coronavirus disease 2019 (COVID-19) hospitalization. A genome-wide association study of expanded mCAs identified 63 significant loci. Germline genetic alleles associated with expanded mCAs were enriched at transcriptional regulatory sites for immune cells. Our results link mCAs with impaired immunity and predisposition to infections. Furthermore, these findings may also have important implications for the ongoing COVID-19 pandemic, particularly in prioritizing individual preventive strategies and evaluating immunization responses.

---

With advancing age comes increased susceptibility to infectious diseases^1,2^. Immunosenescence is the age-related erosion of immune function, particularly with respect to adaptive immunity^12-15^. Leukocytes, including T-cells and B-cells, are key mediators of adaptive host defenses against infections, with impaired immune responses increasing risk for infections^16-18^. Age-related mosaic chromosomal alterations (mCAs) detected from blood-derived DNA, are clonal structural somatic alterations (deletions, duplications, or copy neutral loss of heterozygosity) present in a fraction of peripheral leukocytes that can indicate clonal hematopoiesis (CH)^3-5^. mCAs are associated with aberrant leukocyte cell counts, and increased risks for hematological malignancy and mortality^3-11^.

While the relationship between mCAs and increased hematologic cancer risk is well established^3-5^, the impact of mCAs on age-related diminishment in immune function is poorly understood. We hypothesized that mCAs increase risk of infection since mCAs are somatic variants that increase in abundance with age and are associated with alterations in leukocyte count. In this study, we harnessed DNA genotyping array intensity data and long-range chromosomal phase information inferred from 767,891 individuals across four countries to analyze the associations between expanded mCA clones (i.e., mCAs present in at least 10% of peripheral leukocyte DNA indicative of clonal expansion) and diverse infections, including severe coronavirus disease 2019 (COVID-19) from SARS-CoV-2 infection (**Figure 1a**). To elucidate genetic risk factors for the development of expanded mCA clones, we performed a genome-wide association study (GWAS) in the UK Biobank and subsequent *in silico* cell-specific, transcriptomic, and pathway analyses.

**Figure 1:**
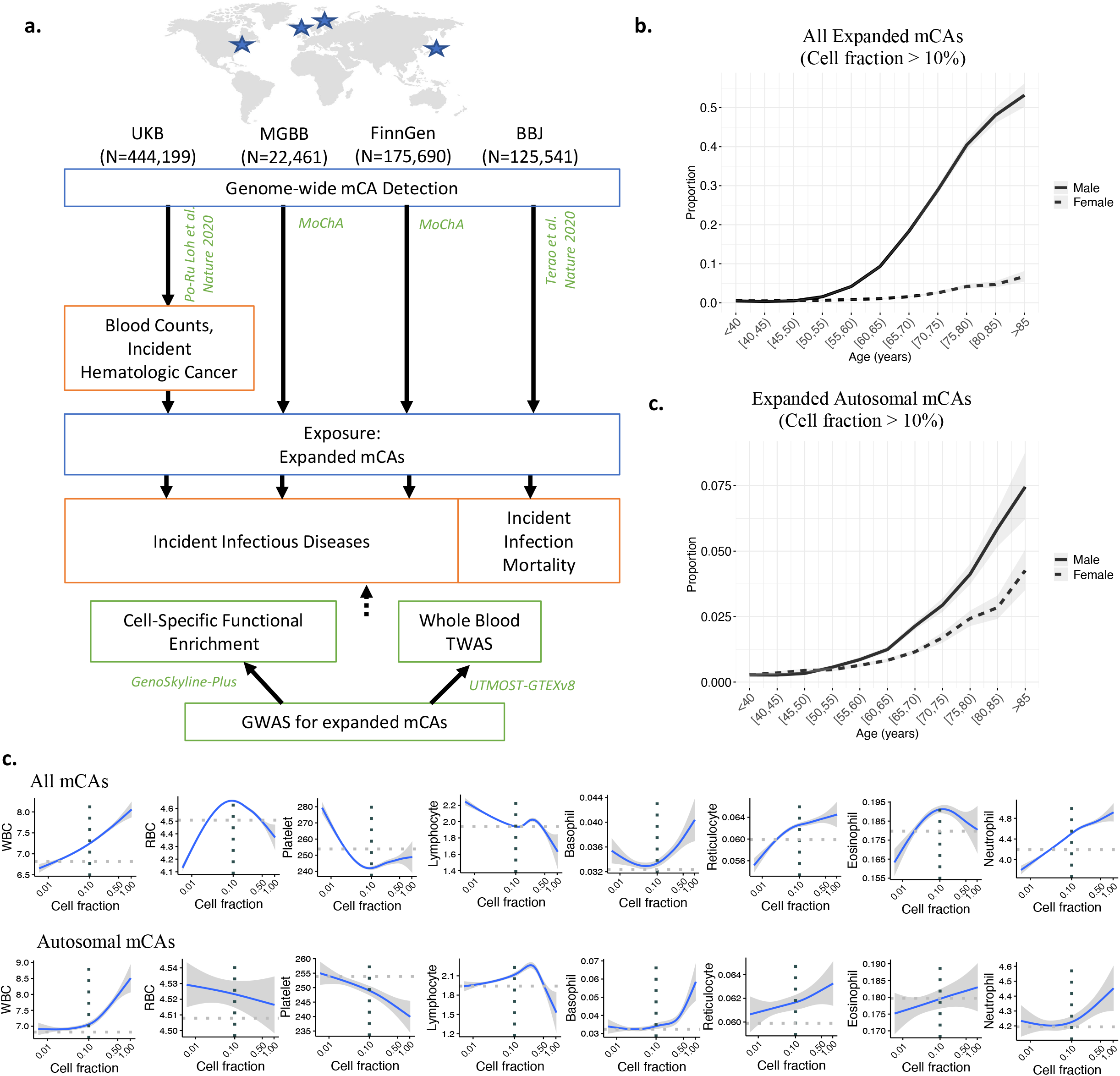
Study schematic. **a**. Genome-wide mCAs were detected across the UKB^4^, MGBB (via the MoChA pipeline), FinnGen (via the MoChA pipeline), and BBJ^3^. Association of expanded mCAs (cell fraction >10%) with incident infectious diseases in UKB, MGBB, and FinnGen and with incident infectious disease mortality in BBJ was performed. A GWAS for expanded mCAs was then performed in the UKB to discover causal factors for expanded mCAs. Using the GWAS results, cell-specific functional enrichment analyses were performed using GenoSkyline-Plus, which combines epigenetic and transcriptomic annotations with GWAS summary statistics to estimate the relative contribution of cell-specific functional markers to the GWAS results. Additionally, to prioritize putative causal genes and pathways promoting the development of expanded mCAs, whole blood TWAS was performed using UTMOST via GTEx v8. Association of **b**. all expanded mCAs with cell fraction >10%, and **c**. all expanded autosomal mCAs, with age using 5-year age bins stratified by sex among individuals in the UKB, MGBB, FinnGen, and BBJ combined. Plots by cohort and across other mCA groupings are available in **Supplementary Note 7, 8. d**. Associations of mCA cell fraction with blood counts (in units of 10^9 cells/L) in the UKB among individuals without prevalent hematologic cancer at time of blood draw for genotyping and cell count measurement. The dotted horizontal lines reflect the mean blood count for individuals without an mCA. The dotted vertical lines at cell fraction of 0.10 represents the cutoff for the expanded mCA definition. Individuals with known hematologic cancer at time of or prior to blood draw for genotyping were excluded. BBJ = BioBank Japan, GTEx v8 = Genotype-Tissue Expression project version 8, GWAS=genome-wide association study, MGBB = Mass General Brigham Biobank, mCA = mosaic chromosomal alterations, MoChA = Mosaic Chromosomal Alterations software (https://github.com/freeseek/mocha), TWAS = transcriptome-wide association study, UKB = UK Biobank, UTMOST = Unified Test for MOlecular SignaTures.

## Results

### Population characteristics and mCA prevalence

A total of 767,891 unrelated, multi-ethnic individuals across the UK Biobank (UKB) (N=444,199), Mass General Brigham Biobank (MGBB) (22,461), FinnGen (N=175,690), and BioBank Japan (BBJ) (N=125,541) passing genotype and mCA quality control criteria (**Supplementary Notes 1-5**) were analyzed (**Supplementary Table 1**). While UKB and BBJ mCA calls were previously performed^3,4^, the MoChA pipeline (https://github.com/freeseek/mocha) was implement to detect mCAs in MGBB and FinnGen (**Extended Data Figure 1**) from genome-wide genotyping of blood DNA in the present study. Among the UKB participants, mean age at DNA collection was 57 (standard deviation [SD] 8) years, 204,579 (46.1%) were male, 188,875 (45.0%) were prior or current smokers, and 66,551 (15.0%) had a history of solid cancer. In the MGBB, mean age was 55 (SD 17) years, 10,306 (45.9%) were male, 9,094 (40.5%) were prior or current smokers, and 6,080 (27.1%) had a history of solid cancer. In FinnGen, mean age was 53 (SD 18) years, 71,000 (40.4%) were male, 42.7% were prior or current smokers (when smoking status was available), and 31,855 (18.1%) had a history of solid cancer. In BBJ, mean age was 65 (SD 12) years, 72,186 (57.5%) were male, and 66,913 (53.3%) were prior or current smokers, and 25,987 (20.7%) had a history of solid cancer.

In the UKB, among 444,199 unrelated individuals without a known history of hematologic malignancy, 66,011 (14.9%) carried an mCA (15,350 autosomal) and 12,398 (3.2%) carried an expanded mCA clone, defined as an mCA mutation present in at least 10% of peripheral leukocytes (2,985 autosomal) (**Supplementary Table 2**). While most of carriers only carried one mCA, 6% of individuals carried between 2 to 22 non-overlapping mCAs (**Supplementary Note 6**). In the MGBB, across 22,461 unrelated individuals without a history of hematologic cancer, 3,784 (16.8%) carried an mCA (1,025 autosomal) and 1,026 (5.2%) carried an expanded mCA clone (337 autosomal). In FinnGen, across 175,690 individuals without a history of hematologic cancer, 22,040 (12.5%) carried an mCA (3,164 autosomal), and 9,558 (5.9%) carried an expanded mCA clone (1,620 autosomal). In BBJ, across 125,541 individuals without a history of hematologic cancer, only autosomal mCAs were available, with 20,440 carriers (16.3%) and 1,676 (1.3%) that carried an expanded clone. (**Supplementary Table 2**).

Consistent with previous reports, the prevalence of mCAs increased with age and was more common among men (**Supplementary Note 7,8**, and **Supplementary Table 3**). Across the UKB, MGBB, FinnGen, and BBJ cohorts combined, the prevalence of expanded mCAs was 0.5% among individuals <40 years, 1.2% among 40-60 years, 7.8% among 60-80 years, and 26.5% among those greater than 80 years (**Figure 1b**), the majority of which is due to loss of X in females and loss of Y in males (**Supplementary Note 7**). The prevalence of expanded autosomal mCAs was 0.27% among individuals <40 years, 0.52% among 40-60 years, 1.5% among 60-80 years, and 4.6% among those greater than 80 years (**Figure 1c**).

### Association of mCAs with hematologic traits

We observed a striking association of mCA cell fraction with aberrant cell blood counts acquired at the same visit as blood for genotyping (**Figure 1d**). Increased mCA cell fraction was associated with overall increased white blood cell count with general consistency across the cell differential components, with inflections at around cell fraction of 0.1 (**Figure 1d**). The strongest association across all mCAs groupings (autosomal/chrX/chrY) with blood counts was between expanded autosomal mCAs and increased lymphocyte count at enrollment (Beta 0.40 SD or 0.25 x10^9^ cells/L; 95% CI 0.36 to 0.44 SD; P=4.2×10^−84^) (**Extended Data Figure 2, Supplementary Note 9**).

Similarly, incident hematologic cancer risk was also strongly dependent on cell fraction (**Extended Data Figure 3a,b**). We reproduced the associations of mCAs with hematologic cancers with similar effects as previously described in the UKB^4,5^. We found that expanded autosomal mCAs with cell fraction >10% were most strongly associated with incident hematologic cancer (**Extended Data Figure 3c**), with the strongest association being for incident chronic lymphocytic leukemia (HR 121.9; 95% CI 93.6 to 158.9; P=4.2×10^−277^); although an association with myeloid leukemia was also present (HR 12.3; 95% CI 7.7 to 19.7; P=2.3×10^−25^) (**Supplementary Figure 11**). While expanded chrX and chrY mCAs were also associated with chronic lymphocytic leukemia, their effects were considerably lower (chrX: HR 24.1, 95% CI 5.8 to 99.9, P=1.1×10^−5^ and chrY: HR 2.0, 95% CI 1.0 to 4.0, P=0.038) (**Extended Data Figure 3c**).

### Associations with diverse infections

mCA presence across the genome was associated with diverse incident infections (defined in **Supplementary Data 1,2**) (HR 1.06; 95% CI 1.04 to 1.09; P=8.6×10^−8^) (**Supplementary Note 10**), independent of age, age^2^, sex, smoking status, and first 10 principal components of ancestry in the combined UKB, MGBB, and FinnGen meta-analysis. The dependence of this association with mCA cell fraction is further visualized in **Figure 2a,b**, which shows an increase in proportion of incident infection cases and incident sepsis cases with cell fraction, with greater slopes observed at approximately cell fraction >10%. Accordingly, the associations across diverse infections were stronger for expanded mCA clones, (HR 1.12; 95% CI 1.1 to 1.2; P=6.3×10^−7^) (**Figure 2c**). Furthermore, among expanded mCA clones, the strongest association was observed among expanded autosomal mCAs (HR 1.3; 95% CI 1.1 to 1.4; P=1.8×10^−7^) (**Figure 2c)**. Accounting for multiple hypothesis testing, expanded autosomal mCAs were significantly associated with sepsis (HR 2.7; 95% CI 2.3 to 3.2; P=3.1×10^−28^), respiratory system infections (HR 1.4; 95% CI 1.2 to 1.5; P=3.8×10^−10^), digestive system infections (HR 1.5; 95% CI 1.3 to 1.7; P=2.2×10^−9^), and genitourinary system infections (HR 1.3; 95% CI 1.1 to 1.4; P=3.7×10^−4^) (**Figure 2c)**. The specific expanded autosomal mCAs implicated for infection were diverse in nature – across all chromosomes, of different sizes, and mixed across gain, loss, and copy-number neutral loss of heterozygosity (CNN-LOH) mCAs (**Extended Data Figure 4**). Further associations across 20 specific infectious disease subcategories are enumerated in **Supplementary Note 11**. For sex chromosome mCAs, none of the incident infections achieved statistical significance (P<0.005) in meta-analysis across the three cohorts; however, respiratory infections were suggestively associated (expanded chrX: HR 1.5; 95% CI 1.01 to 1.9; P=0.0068; expanded chrY: HR 1.09; 95% CI 1.0 to 1.2; P=0.005) (**Supplementary Figure 12)**.

**Figure 2:**
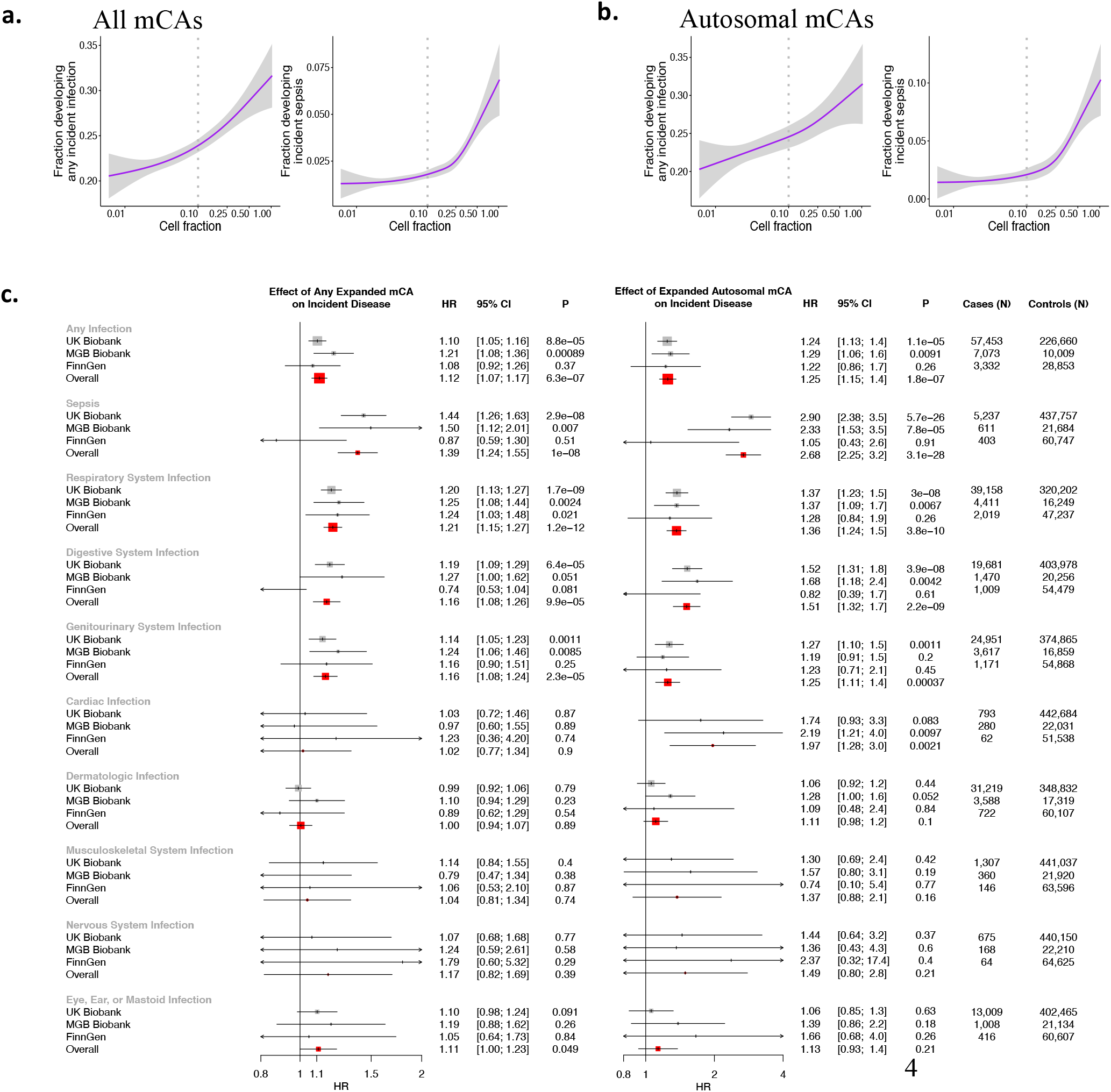
Associations of expanded mCAs with incident infections. Visualizing the dependence with cell fraction among **a**. all mCAs, and **b**. autosomal mCAs, of any incident infection and incident sepsis in the UKB among individuals without prevalent hematologic cancer at time of blood draw for genotyping across. The dotted vertical lines at cell fraction of 0.10 represents the cutoff for the expanded mCA definition. **c**. Association of all expanded mCAs, and separately, expanded autosomal mCAs with incident infections across individuals in the UKB, MGBB, and FinnGen. Analyses are adjusted for age, age^2^, sex, smoking status, and principal components 1-10 of ancestry. Individuals with prevalent hematologic cancer were excluded from analysis. Association analyses for other groupings of mCAs (including across all mCAs regardless of cell fraction, as well as chrX and chrY mCAs are provided in **Supplementary Notes 10, 12**). BBJ = BioBank Japan, MGBB = Mass General Brigham Biobank, mCA = mosaic chromosomal alterations, UKB = UK Biobank.

Risks for incident fatal infections were assessed in BBJ since non-fatal incident infectious disease events are currently unavailable in BBJ. Among individuals without any cancer history in BBJ, autosomal mCAs showed nominal associations with fatal incident infections (HR 1.12, 95% CI 1.0 to 1.2 P=0.04), with expanded autosomal mCAs being associated with incident sepsis mortality (HR 2.0; 95% CI 1.0 to 4.2; P=0.05) (**Supplementary Table 4, Extended Data Figure 5)**, as well as pneumonia history (OR 1.3; 95% CI: 1.1 to 1.5; P=0.0019).

Sensitivity analysis for the association of expanded autosomal mCAs and incident sepsis found that the association was consistently significant across different age groups (**Supplementary Note 13**), and that it was additionally independent of a 25-factor smoking covariate^10^, body mass index, type 2 diabetes mellitus, leukocyte count, lymphocyte count, and lymphocyte percentage (**Supplementary Table 5**).

Stratified analyses indicated expanded autosomal mCAs in individuals with cancer prior to infection (either any solid tumors, or hematologic malignancy after time of blood draw for genotyping) conferred stronger effects for sepsis (HR 2.8; 95% CI 2.3 to 3.4; P=9.7×10^−26^) and respiratory system infections (HR 1.6; 95% CI 1.4 to 1.8; P=6.1×10^−12^) compared to individuals without a prior cancer history (sepsis: HR 1.3; 95% CI 0.8 to 2.0; P=0.33, P_heterogeneity_=0.001; respiratory system infections: HR 1.2; 95% CI 1.0 to 1.3; P=0.045, P_interaction_=0.001) (**Extended Data Figure 6,7; Supplementary Note 14,15**). This interaction was driven by prevalent solid cancer, not hematologic cancer after DNA acquisition for mCA genotyping (**Supplementary Table 6**). Further multivariable adjustment indicated that incident sepsis and infection were independent of chemotherapy, neutropenia, aplastic anemia, decreased white blood cell count, bone marrow or stem cell transplant, and radiation effects prior to infection (with these phenotypes defined using ICD-10 and ICD-9 phecode groupings^19^) (**Extended Table 1**).

### Association with COVID-19 hospitalization

Across 719 COVID-19 hospitalized cases in the UKB, 44 individuals (6%) carried an expanded mCA clone at time of enrollment (in 2010), versus 3% among 337,877 controls. Adjusting for age, age^2^, sex, prior or current smoking status, and principal components of ancestry, expanded mCAs were associated with COVID-19 hospitalizations (OR 1.6; 95% CI 1.1 to 2.2; P=0.0082), with higher effect estimates from expanded autosomal mCAs (OR 2.2; 95% CI 1.2 to 4.1; P=0.02) (**Figure 3a**). Analyses in FinnGen showed evidence of independent replication. The meta-analyzed associations across UKB and FinnGen of expanded autosomal mCAs on COVID-19 hospitalization was OR 2.4, 95% CI 1.3 to 4.5, P=0.004 (**Figure 3a,b**). Similar to prior phenotypes, the fraction of individuals hospitalized with COVID-19 increased with cell fraction, with particularly strong slopes after cell fraction >10% (**Figure 3c**). In the UKB, further sensitivity analysis was performed; the associations persisted with additional adjustment for normalized Townsend deprivation index, normalized body mass index, type 2 diabetes mellitus, hypertension, coronary artery disease, any cancer, asthma, and chronic obstructive pulmonary disease (**Extended Data Figure 8a**). Additionally, similar associations were observed in the UKB when comparing COVID-19 hospitalization to tested negative controls, COVID-19 positive to all from English provinces and, COVID-19 positive to tested negative controls (**Extended Data Figure 8b**). Similar to the diverse nature of mCA clones observed in cases of incident infection, specific mCA clones carried by COVID-19 hospitalized individuals were also diverse in nature – across multiple chromosomes, a wide range of sizes, and both gain, loss, and CNN-LOH copy changes (**Figure 3d**). Similar effects associations effects of expanded mCAs with COVID-19 were also observed with incident pneumonia in the UKB (**Extended Data Figure 8c**).

**Figure 3:**
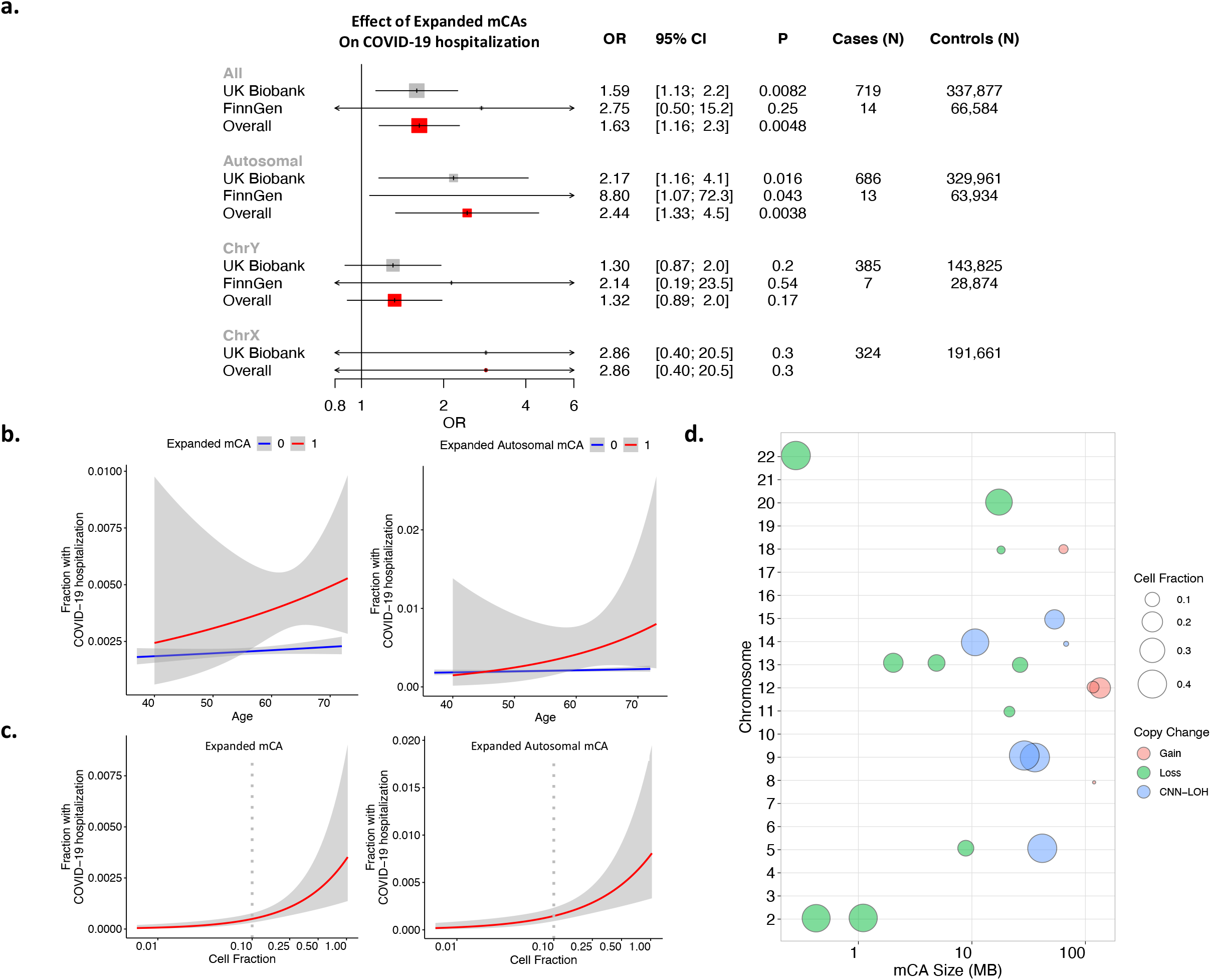
Association of expanded mCAs with COVID-19 Hospitalization. **a**. Associaiton of expanded mCAs with COVID-19 Hospitalization across the UKB and FinnGen. Individuals with known hematologic cancer at time of or prior to blood draw for genotyping were excluded. Analyses are adjusted for age, age^2^, sex, ever smoking status, and principal components of ancestry. b. Fraction of COVID-19 hospitalizations plotted by age, stratified by Expanded mCA (left) and expanded autosomal mCA (right) c. Fraction of COVID-19 hospitalizations plotted by cell fraction among expanded mCAs (left) and expanded autosomal mCAs (right). **d**.Visualization of the diverse range of expanded autosomal mCAs detected across the genome among individuals hospitalized with COVID-19 in the UK Biobank. Each point represents one mCA carried by a case, with the x-axis as the chromosome, y-axis as the mCA size in mega-bases of DNA (MB). Additional sensitivity analyses in the UKB are provided in **Extended Data Figure 8**. MGBB = Mass General Brigham Biobank, UKB = UK Biobank, MB=megabase, CNN-LOH = copy number neutral loss of heterozygosity

### Germline genetic predisposition to expanded mCAs

To further elucidate causal factors for expanded mCA clones, we performed a genome-wide association study (GWAS) in the UKB. We identified 63 independent genome-wide significant loci (r^2^< 0.1 across 1MB windows of the genome) (**Figure 4a, Supplementary Data 3**). Across the 63 germline variants, significant correlation was seen between different mCA categories (**Supplementary Note 16**), suggesting the presence of shared germline genetic variants predisposing to mCAs across the genome. Follow-up analyses using an additive polygenic risk score comprised of 156 independent genome-wide significant variants associated with mosaic loss-of-chromosome Y (mLOY) from males from a prior study in the UKB^20^, found significant associations with expanded autosomal mCAs and expanded ChrX mCAs in females, further highlighting the shared germline contributors towards mCAs across the genome (**Supplementary Note 17**). TWAS combining the expanded mCA GWAS results with GTEXv8^21^ whole blood expression quantitative trait loci (eQTLs) using UTMOST^22^ prioritized 62 genes (P<3.2×10^−6^) promoting expanded mCA development (**Figure 4b**). While gene enrichment analyses with the Elsevier Pathway Collection did not identify significantly associated pathways after multiple testing correction, top pathways were linked to DNA damage repair and lymphoid processes (**Extended Data Figure 9a**). The corresponding GWAS locus-zoom plots for some of these immune-related genes are shown in **Extended Data Figure 9b**. To prioritize tissues most implicated by these loci, tissue enrichment analyses using GenoSkyline-Plus were performed. Significant enrichment was identified in immune-specific epigenetic and transcriptomic functional regions of the genome (P=7.1×10^−9^) (**Figure 4c**). Further stratification of the immune category identified specific enrichment for CD4+ T-cells (P=0.00098) (**Figure 4d**).

**Figure 4:**
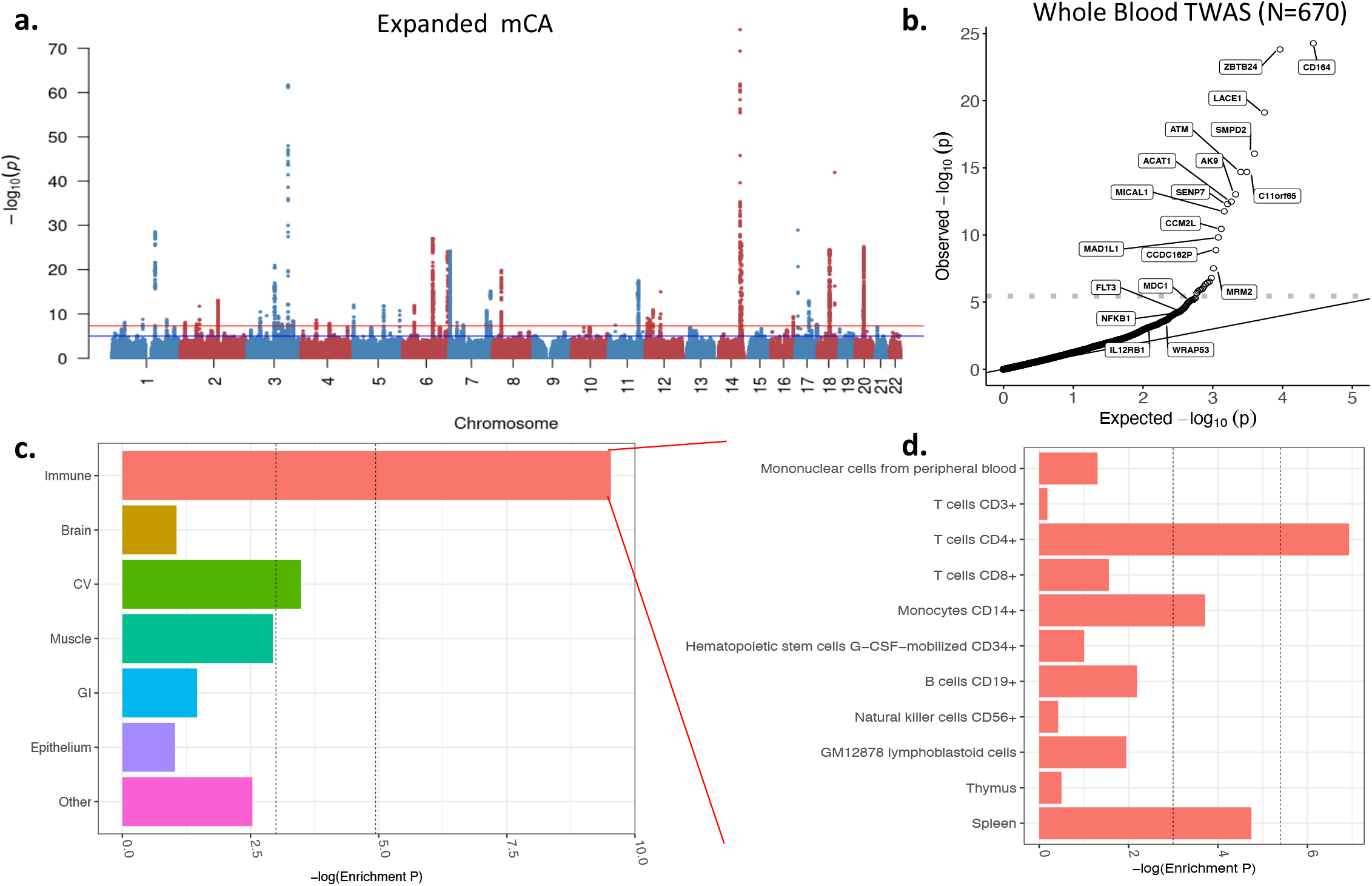
Inherited risk factors for expanded mCAs: **a**. GWAS, TWAS, and Cell Type Enrichment. GWAS for expanded mCA identified 63 independent loci. **b**. Quantile-quantile plot of the whole blood TWAS of the expanded mCA GWAS using 670 samples from GTExv8 shows enrichment across 62 genes. The horizontal dotted line reflects the Bonferroni-adjusted p-value for significance. Genes with TWAS P<5×10^−8^ or those important in the pathway-enrichment analyses from **Extended Data Figure 9** are labeled. c. cell-type enrichment results from the Expanded mCA GWAS across immune, brain, cardiovascular (CV), muscle, gastrointestinal (GI), epithelium, and other tissues as annotated using GenoSkyline-Plus annotations. D. Zooming in to show the stratified enrichment by specific categories of immune cells and tissues. Across panels C. and D., the vertical dotted lines indicate (1) P=0.05 for suggestive enrichment, and (2) the Bonferroni-adjusted P-value for significant enrichment. GWAS = genome wide association study, TWAS = transcriptome-wide association study, CV = cardiovascular, GI = Gastrointestinal

## Discussion

Across four geographically distinct biobanks comprising 767,891 individuals without known hematologic malignancy, clonal hematopoiesis (CH) represented by expanded mCAs is increasingly prevalent with age but not readily detectable by conventional medical blood tests. In addition to strongly predicting future risk of hematologic malignancy, expanded mCAs were also associated with risk for diverse incident infections, particularly sepsis and respiratory infections. These findings were robust across age, sex, tobacco smoking, and were strongest among those who develop cancer. Consistent with these observations, expanded mCAs were also associated with increased odds for COVID-19 hospitalization.

These results support several conclusions. First, mCA-driven CH is a potential risk factor for infection. Recent work showed that CH with myeloid malignancy driver mutations, also referred to as ‘clonal hematopoiesis of indeterminate potential’ (CHIP), predisposes to myeloid malignancy and coronary artery disease^23-27^. Meanwhile, CH with larger chromosomal alterations (i.e., mCAs) predisposes primarily to lymphoid malignancy but not coronary artery disease^3-5,8,9^. Our observations suggest CH defined by the presence of mCAs is a risk factor for infection. Since the relationship between mCAs and infection risk was not substantially attenuated when adjusting for leukocyte or lymphocyte counts at baseline visit, the impact of mCAs on infection risk likely acts through mechanisms independent of the impact of CH on cell counts. For example, as mCAs alter gene dosage (e.g., via duplications and deletions) and remove allelic heterogeneity (e.g., copy neutral loss-of-heterozygosity events) in leukocytes, potential impacts on the differentiation, function, and survival of leukocytes are mechanisms that could lead to altered infection risk. Our germline analyses specifically implicate lymphoid tissues. In particular, many of the mCA susceptibility loci are the same as those found in chronic lymphocytic leukemia, a condition in which lymphocyte differentiation and function is altered promoting infection risk^28-31^. Therefore, molecular changes in leukocytes that promote clonal expansion may occur at the expense of reduced ability to combat infection.

Second, the infectious disease risk associated with mCAs is exacerbated in the setting of cancer. It is well-established that mCAs in blood-derived DNA increase risk for hematologic cancer^3-5^. Furthermore, recent evidence suggests an association between mCAs detected in blood-derived DNA and increased risk of select solid tumor^7,10,32^. Our analysis identified an interaction between mCAs and prior cancer diagnosis that amplified sepsis and pneumonia risk. Importantly, this interaction was restricted to individuals with solid cancers, not antecedent blood cancer. While this observation could be partially due to synergistic immunosuppressive side effects of cancer therapies^33^, the observed associations persisted despite adjustment for many of these treatments. Alternatively, abnormal regulation of immune inflammatory pathways that release cytokines and inflammatory cells may create chronic states of inflammation in individuals with mCAs^34,35^. Surveillance for expanded mCA clones, particularly among those who develop solid cancer, may help identify individuals at high risk for infection that could benefit from targeted interventions.

Third, our findings could have particular relevance for the ongoing COVID-19 pandemic. We observed that mCAs are associated with elevated risk for COVID-19 hospitalization, with greater than two-fold risk linked to expanded autosomal mCAs. Maladaptive immune responses, particularly in leukocytes, increase risk for severe COVID-19 infections^36-39^. Awareness of COVID-19 risk associated with mCAs may help with the prioritization of emerging prophylactic treatments and initial vaccination programs. However, whether immune response to conventional vaccination approaches is altered in the context of mCAs deserves further study.

This analysis of mCAs and infection had some limitations. First, our study only measures mCAs at one time point for each participant. While our sampled mCA time point is likely correlated with CH at time of infection, CH dynamically changes over time potentially leading to differences in cellular fraction or additional undetected events that were acquired prior to infection. Second, we cannot rule out the possibility of undiagnosed hematologic malignancy among individuals with mCAs with only blood DNA. However, given the observed prevalence of mCAs (4% by age 60 years) among individuals without diagnosed hematologic malignancy and general scarcity of hematologic malignancy in the general population, we anticipate undiagnosed hematologic malignancy at DNA acquisition to be uncommon. Third, despite the robust adjustment and sensitivity analyses performed in our statistical analysis, including adjustment for chemotherapy, bone marrow transplant, radiation, and other features associated with poor cancer prognosis (neutropenia, aplastic anemia, decreased white blood cell count), we cannot completely rule out the impact of residual confounding in our results from unknown or unmeasured sources. Consistency across cohorts and infection types and biologic plausibility mitigates this possibility, but functional studies testing the hypothesis that these represent causal relationships merit consideration.

In conclusion, we report evidence for increased susceptibility to a spectrum of infectious diseases in individuals carrying mCAs in a detectable fraction of leukocytes. The impacts of mCA on infection risk are systemic, with increased susceptibility to infection observed for a variety of organ systems, including severe COVID-19 presentations.

## Online Methods

### Study samples

A total of 767,891 individuals across four biobanks were analyzed: UK Biobank (UKB), Mass General Brigham Biobank (MGBB), FinnGen, and Biobank Japan (BBJ)^40-42^. Across all three cohorts, written informed consent was previously obtained from all participants. Individuals with known hematologic cancer at time of or prior to blood draw for genotyping were removed from all analyses. Additional information on each cohort is provided in **Supplementary Note 1**.

### Mosaic chromosomal alteration detection

Mosaic chromosomal alteration (mCA) detection has been previously described in the UKB^4,5^ and BBJ^3^. mCA detection in the MGBB and FinnGen were performed with the Mosaic Chromosomal Alterations (MoChA) software and pipeline (https://github.com/freeseek/mocha). Briefly, genotype intensities were transformed to log_2_(R ratio) (LRR) and B-allele frequency (BAF) values to estimate total and relative allelic intensities, respectively, as previously described^43^. Further details regarding the mCA detection are provided in **Supplementary Note 1-5**. Across all three studies, expanded mCA refers to the presence of at least one detectable mCA present in >10% of circulating leukocytes (e.g., cell fraction >10%). A 10% cell fraction threshold was employed since this has been previously linked to greater clonal haematopoiesis-related risk for incident mortality^10^ and myocardial infarction^23^, additionally this subset was observed to most strongly associate with phenotypes in the UK Biobank including aberrant blood cell counts, incident hematologic cancer, and incident infections (**Figure 1d, 2a,b, Extended Data Figure 3ab**). Autosomes and sex chromosomes were also separately considered; only autosomal mCAs were available for BBJ.

### Clinical outcomes

Definitions for infection outcomes are detailed in **Supplementary Data 1,2**. In the UKB, the first reported occurrences over median 8-year follow-up in Category 2410 were used as categorized by the UKB which maps primary care data, ICD-9 and ICD-10 codes from hospital inpatient data, ICD-10 codes in death register records, and self-reported medical conditions reported at the baseline, to ICD-10 codes. For each set of phenotypes grouped by organ system or by category, the time to first incident event after baseline examination in individuals free of prevalent history of each disease category was used. In the MGBB, electronic health record data was used to define incident ICD-10 codes grouped in the same fashion after DNA collection date over a median 3-year follow-up. In FinnGen, phenotypes were grouped together across ICD-8, ICD-9, and ICD-10 codes (**Supplementary Data 2**), with incident infections defined after DNA collection date over a median 3-year follow-up. In BBJ, analyses were performed using fatal incident events attributed to diverse infection outcomes in **Supplementary Data 1** since non-fatal incident events were not available; additionally, analyses for pneumonia were performed using history of pneumonia prior to genotyping, based on interviews and medical record reviews^41^. Other clinical phenotypes defined in the UKB, MGBB, and FinnGen are detailed in **Supplementary Note 1** and **Supplementary Data 6-8**.

UKB coronavirus disease 2019 (COVID-19), from SARS-CoV-2 infection, phenotypes used in the present analysis were downloaded on July 27, 2020. SARS-CoV-2 infection was determined by polymerase chain reaction from nasopharygeal, oropharyngeal, or lower respiratory samples obtained between March 16, 2020 and July 17, 2020. COVID-19 hospitalized cases were defined as any individual with at least one positive test who also had evidence for inpatient hospitalization (Field 40100). Controls included two sets: (1) participants from UKB English recruitment centers who were not known to have COVID-19, which were individuals with negative or no known SARS-CoV-2 testing or (2) participants with a negative SARS-CoV-2 test. Individuals with COVID-19 of unknown or low severity (i.e., at least one positive SARS-CoV-2 test without a known hospitalization) were excluded from the primary analyses. Replication was performed in FinnGen where SARS-CoV-2 infection was determined either by polymerase chain reaction or by antibodies for samples obtained between March 2, 2020 and July 27, 2020. Across both cohorts, individuals who died prior to March 1, 2020, and therefore were not at risk for COVID-19 infection, were excluded from COVID-19 analyses.

### Statistical methods for infection associations

Association analyses of expanded mCAs with primary incident infection across 10 main infectious disease organ system categories (listed under “organ system” in **Supplementary Data 1**) were performed using Cox proportional hazards models, adjusting for age, age^2^, sex, ever smoking status, and principal components 1-10 from the genotyping data. Time since DNA collection was used as the underlying timescale. The proportional hazards assumption was assessed by Schoenfeld residuals and was not rejected. Individuals with a history of hematological cancer prior to DNA collection were excluded. P-value threshold for significance among the primary organ system infection analyses was two-sided 0.05/10=0.005 to account for multiple hypothesis-testing. Secondary and sensitivity analyses are detailed in the **Supplementary Note 1**. Analyses of incident events were performed separately in each biobank using the survival package in R (version 3.5, R Foundation, Vienna, Austria). Meta-analyses of the UKB, MGBB, and FinnGen results were performed using a fixed effects model from the meta package.

For UKB COVID-19 analyses, logistic regression was performed to estimate the association between expanded mCAs and COVID-19 hospitalization using the aforementioned phenotype definition, adjusting for sex, age, age^2^, smoking status, and the first ten principal components from the genotyping data. As above, individuals with prevalent hematologic cancer were excluded from analyses. For the COVID-19 analyses, statistical significance was assigned at two-sided p-value < 0.05. Secondary multi-variable models were additionally adjusted for normalized Townsend deprivation index^44^, inverse rank normalized body mass index at baseline, type 2 diabetes mellitus, hypertension, coronary artery disease, any cancer, asthma, and chronic obstructive pulmonary disease.

### Genome-wide association study

GWAS was performed using Hail-0.2 software (https://hail.is/) on the Google cloud. Variants were filtered to high-quality imputed variants (INFO score >0.4), with minor allele frequency >0.005, and with Hardy-Weinberg Equilibrium P>1×10^−10^, as previously performed. A Wald-logistic regression model was used for analysis, adjusting for age, age^2^, sex, ever smoking, PC1-10, and genotyping array. Significant, independent loci were identified using P<5×10^−8^ and clumping in Plink-2.0 using an r^2^ threshold of 0.1 across 1MB genomic windows using the 1000-Genomes Project European reference panel. An additive mLOY polygenic risk score was developed as such: 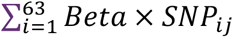, where *Beta* is the weight for each of the 156 independent genome-wide significant variants previously identified in UKB males^20^ and *SNP*_*ij*_ is the number of alleles (i.e., 0, 1, or 2) for *SNP*_*i*_ in female *j* in the UKB.

### Cell-type enrichment analyses

We applied partitioned LD score regression using the LDSC software^45^ to perform enrichment analysis using the expanded mCA GWAS summary statistics in combination with tissue-specific epigenetic and transcriptomic functionality annotations from GenoSkyline-Plus^22^. In addition to the baseline annotations for diverse genomic features as suggested in the LDSC user manual, we specifically examined the enrichment signals on two tiers of annotations of different resolutions: GenoSkyline-Plus functionality scores of 7 broad tissue clusters (immune, brain, cardiovascular, muscle, gastrointestinal tract, epithelial, and others); and GenoSkyline-Plus functionality scores of 11 tissue and cell types within the immune cluster (listed in **Figure 8D**).

### Transcriptome-wide association and pathway enrichment analysis

Transcriptome-wide association was performed using the expanded mCA GWAS summary statistics in combination with the UTMOST^46^ whole blood model updated to GTEXv8 (N=670). Significant genes were identified using a Bonferroni cutoff of P<0.05/15,625 or 3.2×10^−6^. Pathway enrichment analyses was performed using genes with TWAS P<0.001 using the Elsevier Pathways through the EnrichR web server^47^.

### Data Availability

UKB individual-level data are available for request by application (https://www.ukbiobank.ac.uk). The mCA call set was previously returned to the UK Biobank (Return 2062) to enable individual-level linkage to approved UK Biobank applications. Individual-level MGBB data are available from https://personalizedmedicine.partners.org/Biobank/Default.aspx, but restrictions apply to the availability of these data, which were used under IRB approval for the current study, and so are not publicly available. The BBJ genotype data is available from the Japanese Genotype-phenotype Archive (JGA; http://trace.ddbj.nig.ac.jp/jga/index_e.html) under accession code JGAD00000000123. Individual-level linkage of mosaic events can be provided by the BBJ project upon request (https://biobankjp.org/english/index.html). FinnGen data may be accessed through Finnish Biobanks’ FinnBB portal (www.finbb.fi). Additionally, the full expanded mCA genome wide association summary statistics have been uploaded onto the LocusZoom website (https://my.locuszoom.org/gwas/525823/). The present article includes all other data generated or analyzed during this study.

### Code Availability

A standalone software implementation (MoChA) of the algorithm used to call mCAs is available at https://github.com/freeseek/mocha. A pipeline to execute the whole workflow from raw files all the way to final mCA calls is available in WDL format for the Cromwell execution engine as part of MoChA. Code for all other computations are available upon request from the corresponding authors.

## Data Availability

UKB individual-level data are available for request by application (https://www.ukbiobank.ac.uk). The mCA call set was previously returned to the UK Biobank (Return 2062) to enable individual-level linkage to approved UK Biobank applications. Individual-level MGBB data are available from https://personalizedmedicine.partners.org/Biobank/Default.aspx but restrictions apply to the availability of these data which were used under IRB approval for the current study and are not publicly available. The BBJ genotype data is available from the Japanese Genotype-phenotype Archive (http://trace.ddbj.nig.ac.jp/jga/index_e.html) under accession code JGAD00000000123. Individual-level linkage of mosaic events can be provided by the BBJ project upon request (https://biobankjp.org/english/index.html). FinnGen data may be accessed through Finnish Biobanks FinnBB portal (www.finbb.fi). Additionally the full expanded mCA genome wide association summary statistics have been uploaded onto the LocusZoom website (https://my.locuszoom.org/gwas/525823/). The present article includes all other data generated or analyzed during this study.

## Acknowledgements

Thanks to Chris Whelan, Chris Llanwarne, Jason Cerrato, Kyle Vernest, and Khalid Shakir and many other members of the Terra/Cromwell team for their help and advice in the development of the MoChA pipeline. Thanks to Petr Danecek for implementing critical features needed in BCFtools. Thanks to Stephen Chanock for critical input and comments. Thanks to Erikka Loftfield for assistance with the 25-level smoking adjustment variable. Thanks to the participants and staff of the UKB, MGBB, and BBJ. UKB analyses were conducted using Applications 7089 and 21552.

---

**Extended Data Figure 1:**
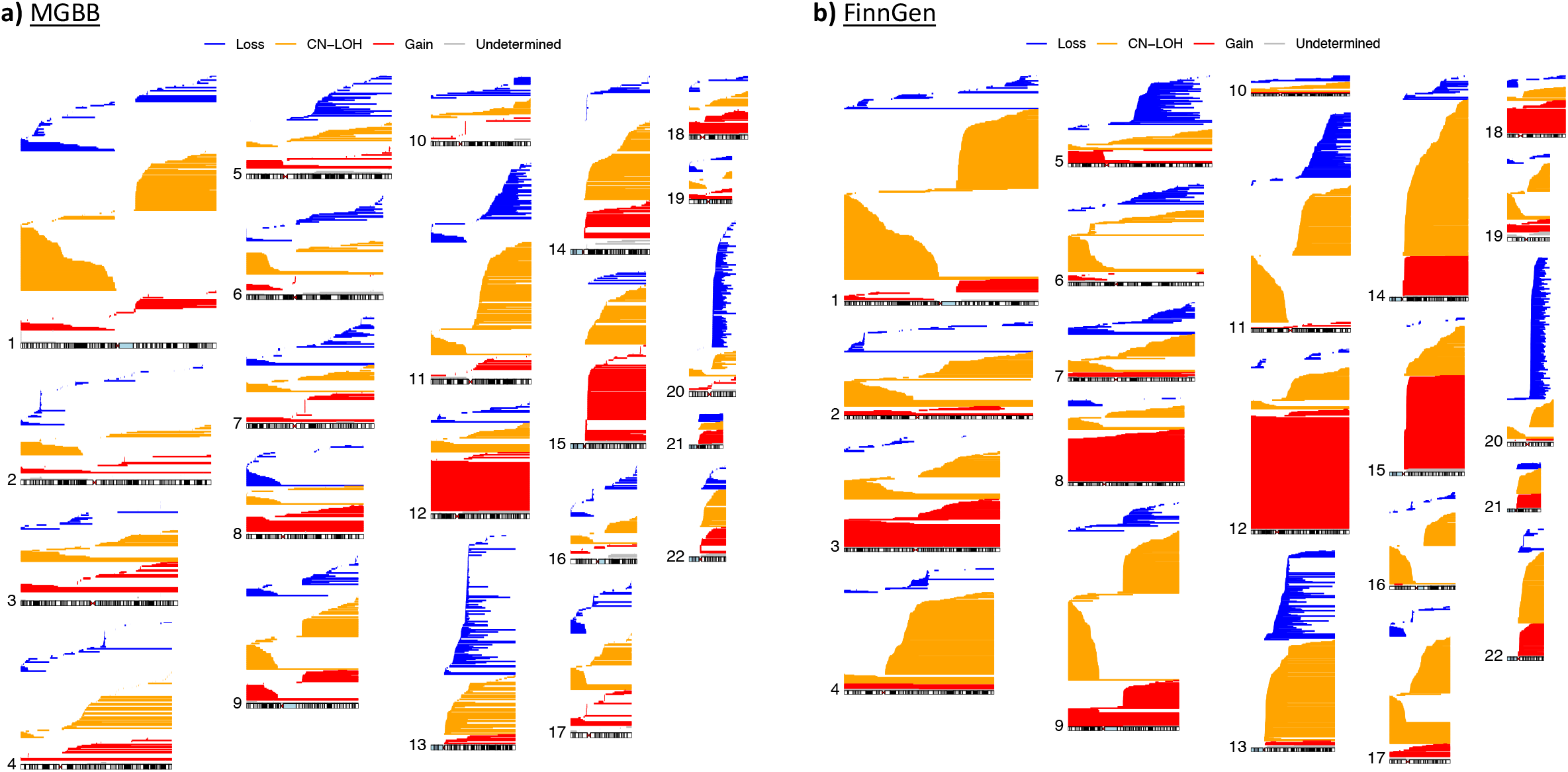
mCA calls by chromosome in the MGBB and FinnGen, CN-LOH = copy neutral loss of heterozygosity

**Extended Data Figure 2:**
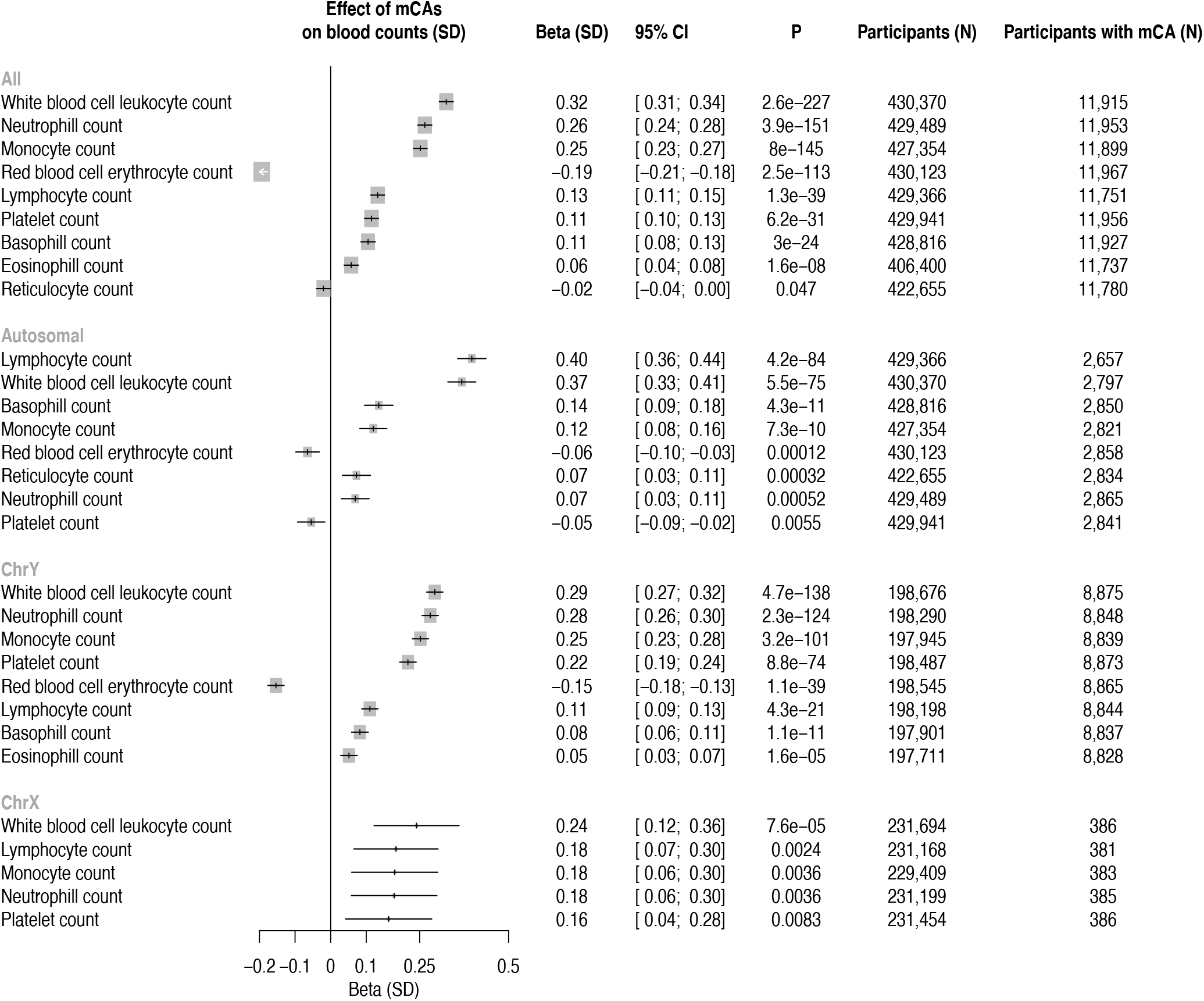
Association of blood counts with expanded mCAs. Associations are adjusted for age, age^2^, sex, smoking status, and principal components of ancestry. mCA = mosaic chromosomal alterations.

**Extended Data Figure 3:**
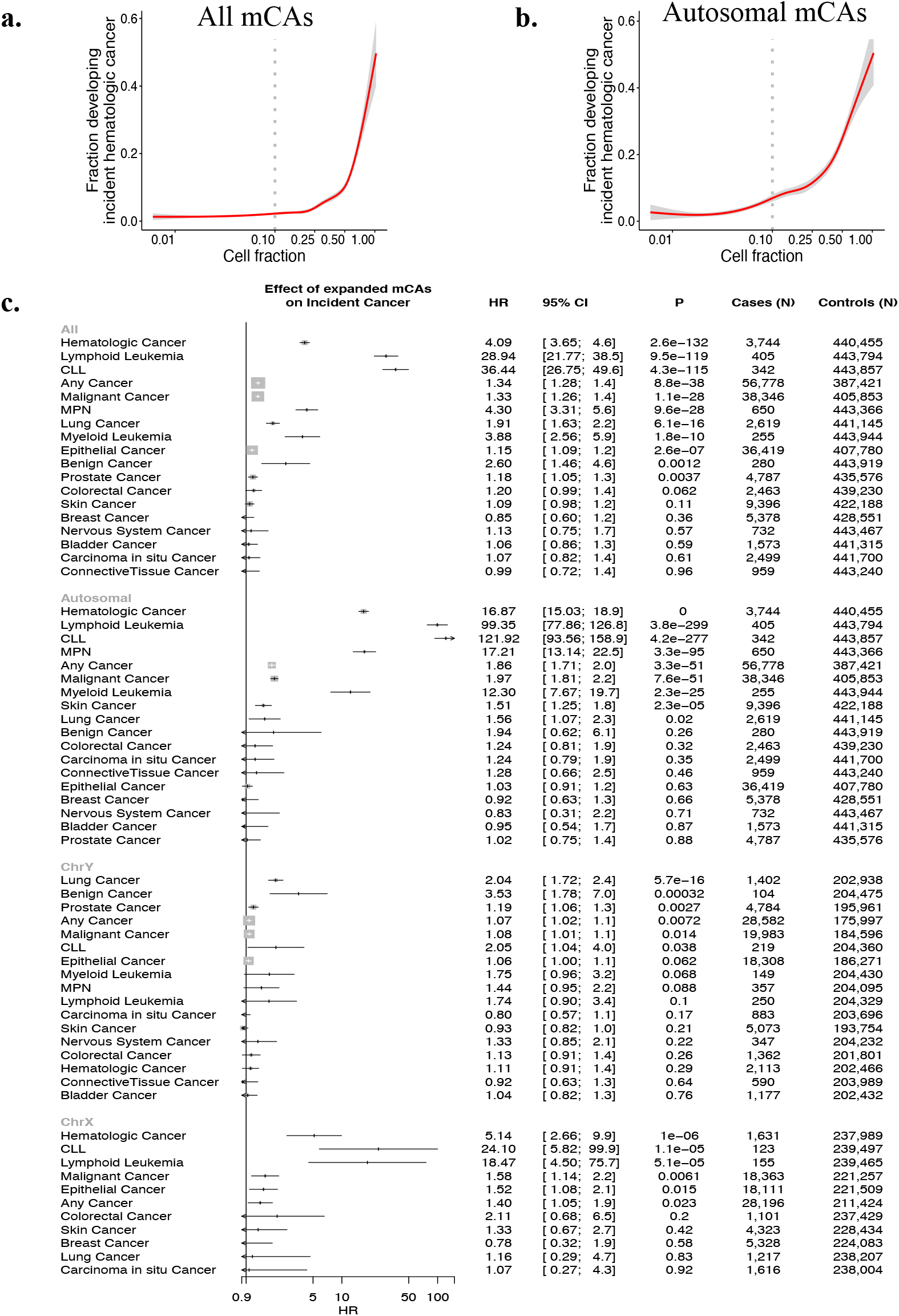
Association of mCAs with incident cancer in the UK Biobank. Association of a) all mCA and b) autosomal mCA cell fraction with incident hematologic cancer. The dotted vertical line at cell fraction of 0.1 shows the cutoff point for expanded mCAs (defined as mCAs with cell fraction >10%). c) Association of expanded mCA categories (with cell fraction>10%) with incident cancer in the UK Biobank. Analyses are adjusted for age, age^2^, sex, smoking status, and principal components of ancestry. Individuals with a history of hematologic cancer at enrollment were removed from analysis. CLL = chronic lymphocytic leukemia, MPN = myeloproliforative neoplasm, mCA = mosaic chromosomal alterations

**Extended Data Figure 4:**
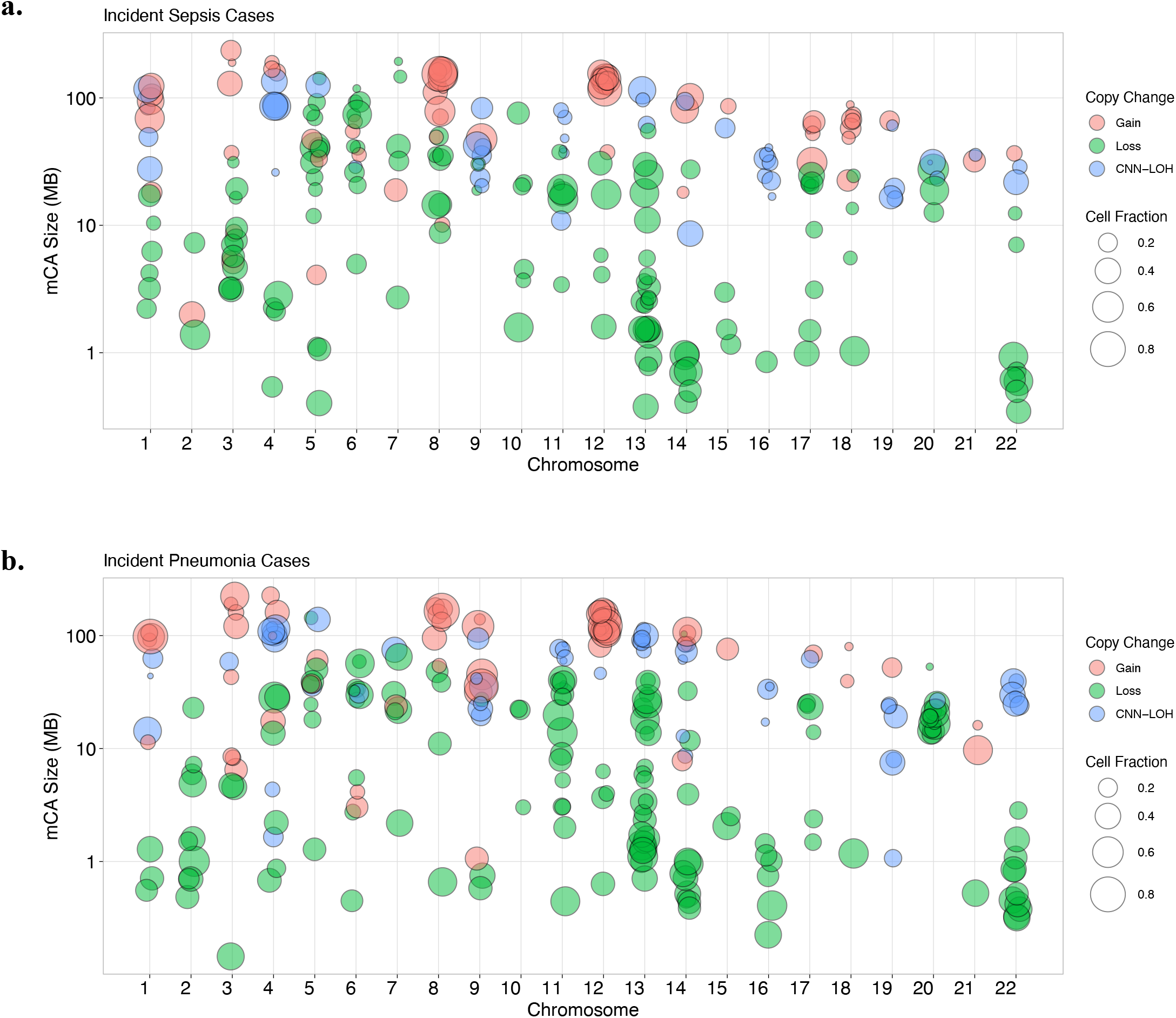
Visualization of the diverse range of expanded autosomal mCAs detected across the genome among individuals with a. incident sepsis and b. incident pneumonia in the UKB. Each point represents one mCA carried by a case, with the x-axis as the chromosome, y-axis as the mCA size in mega-bases of DNA (MB), color as the copy change, and size of the point as the cell fraction of that mCA. CNN-LOH=copy number neutral loss of heterozygosity, MB = megabases of DNA, mCA = mosaic chromosomal alterations

**Extended Data Figure 5:**
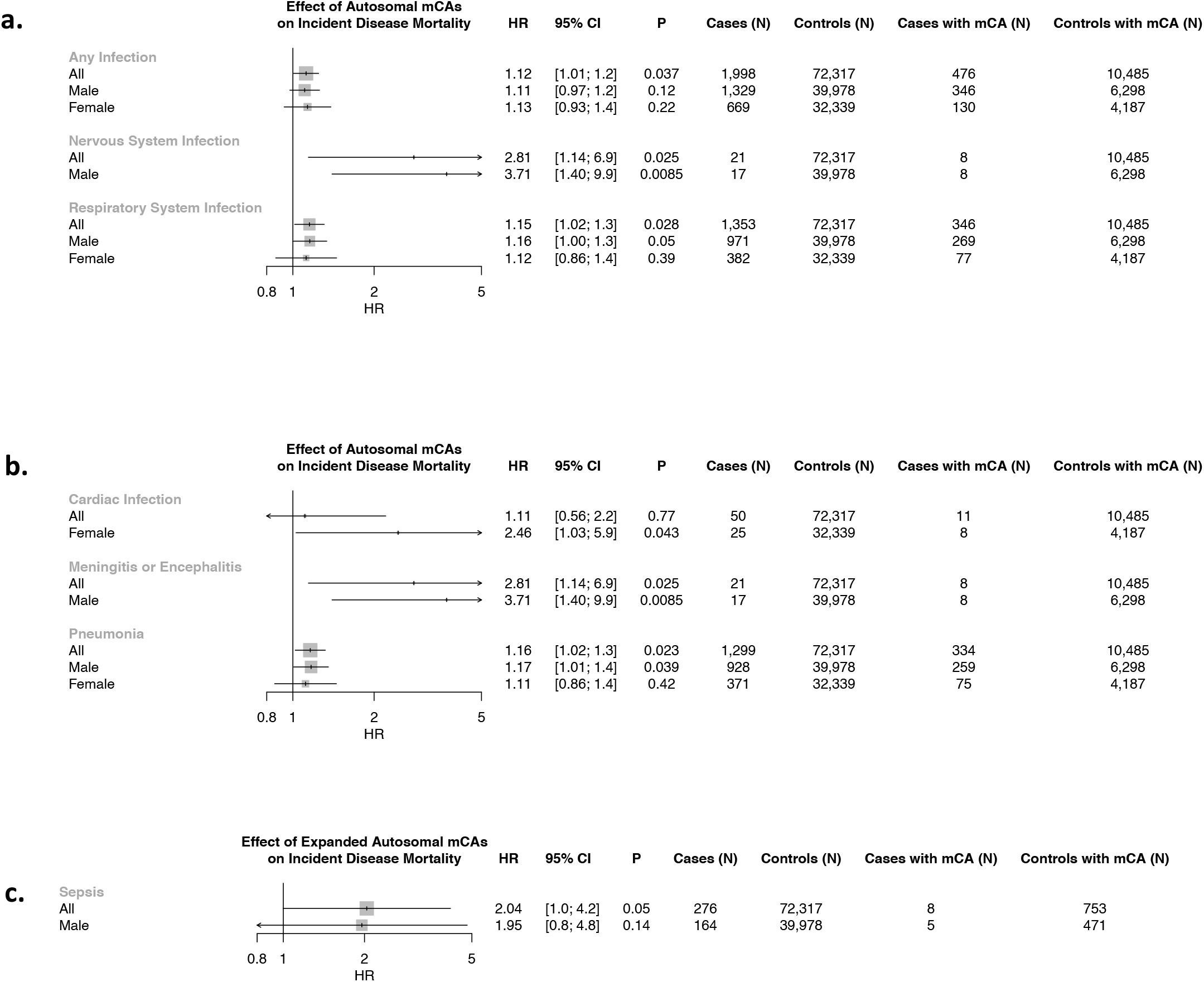
Suggestive associations (P<0.05) of mCAs with incident infection-related mortality in Biobank Japan. Associations of autosomal mCAs with a) organ-system level infections and b) specific infection categories. c) Association of expanded autosomal mCAs with Sepsis. Full results are in Supplementary Table 6. Associations are presented among individuals without any cancer history. mCA = mosaic chromosomal alterations.

**Extended Data Figure 6:**
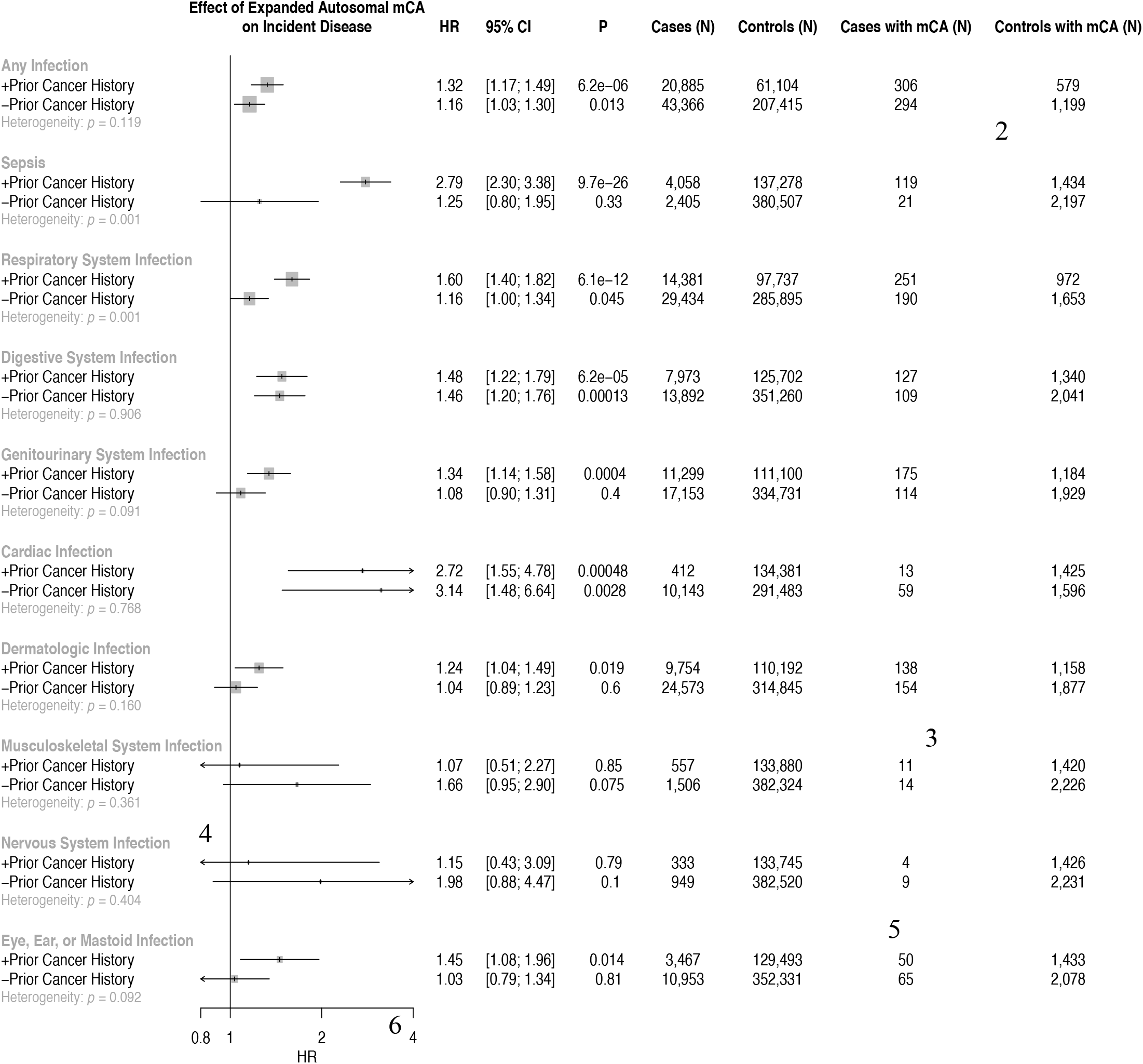
Association of expanded autosomal mCAs with incident infections across individuals with and without a cancer history before their incident infection, meta-analyzed across UKB, MGBB, and FinnGen combined (cohort-specific analyses are available in Supplementary Figure 15). Individuals with known hematologic cancer at time of or prior to blood draw for genotyping were excluded. Analyses are adjusted for age, age2, sex, smoking status, and principal components of ancestry. mCA = Mosaic chromosomal alteration, MGBB = Mass General Brigham Biobank, UKB = UK Biobank

**Extended Data Figure 7:**
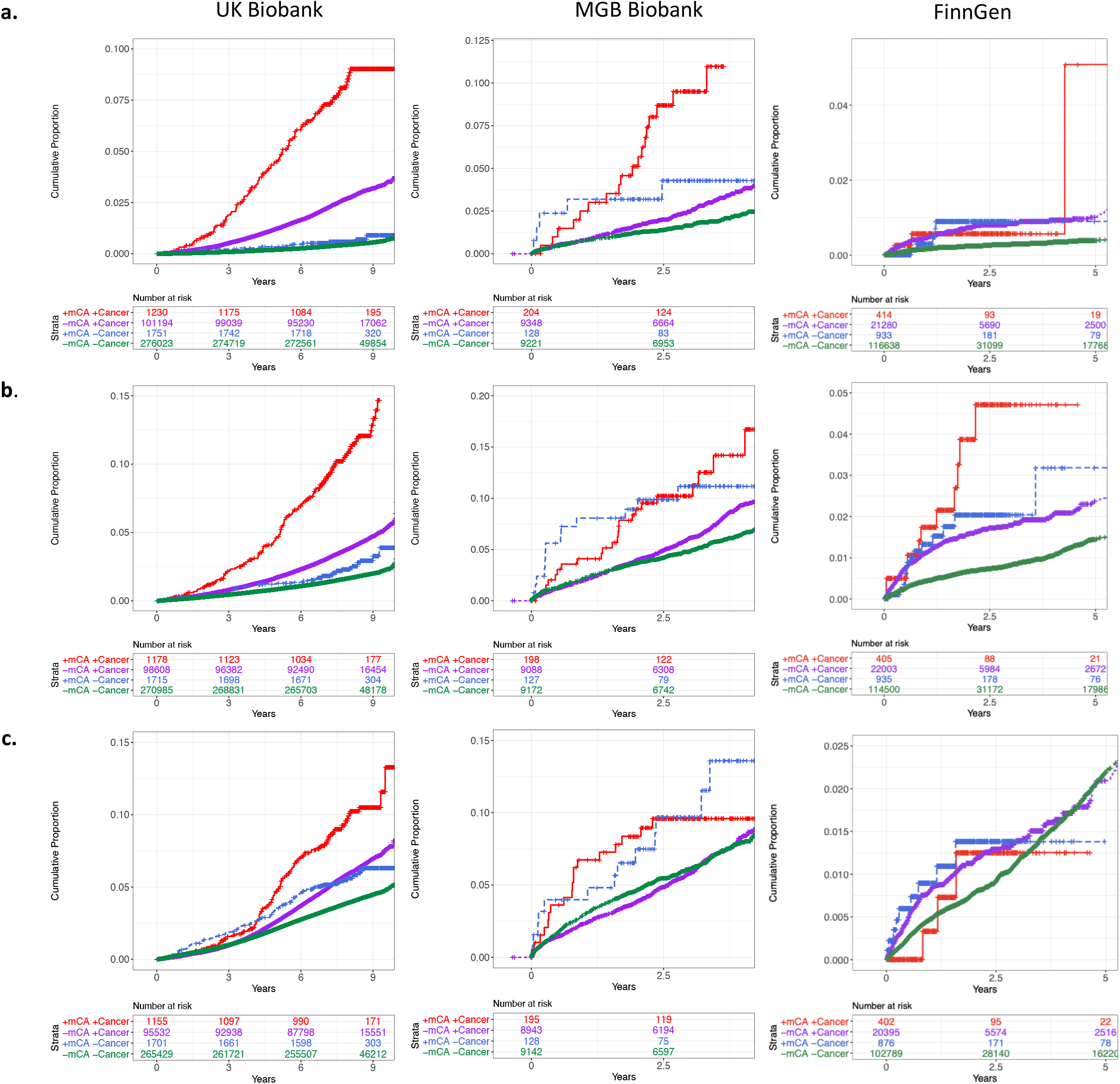
Association of expanded autosomal mCAs with incident **a**. sepsis, **b**. pneumonia, and **c**. digestive system infection across carrier status for expanded autosomal mCAs and any cancer diagnosis prior to the incident infection date. Individuals with known hematologic cancer at time of or prior to blood draw for genotyping were excluded. mCA = mosaic chromosomal alterations.

**Extended Data Figure 8:**
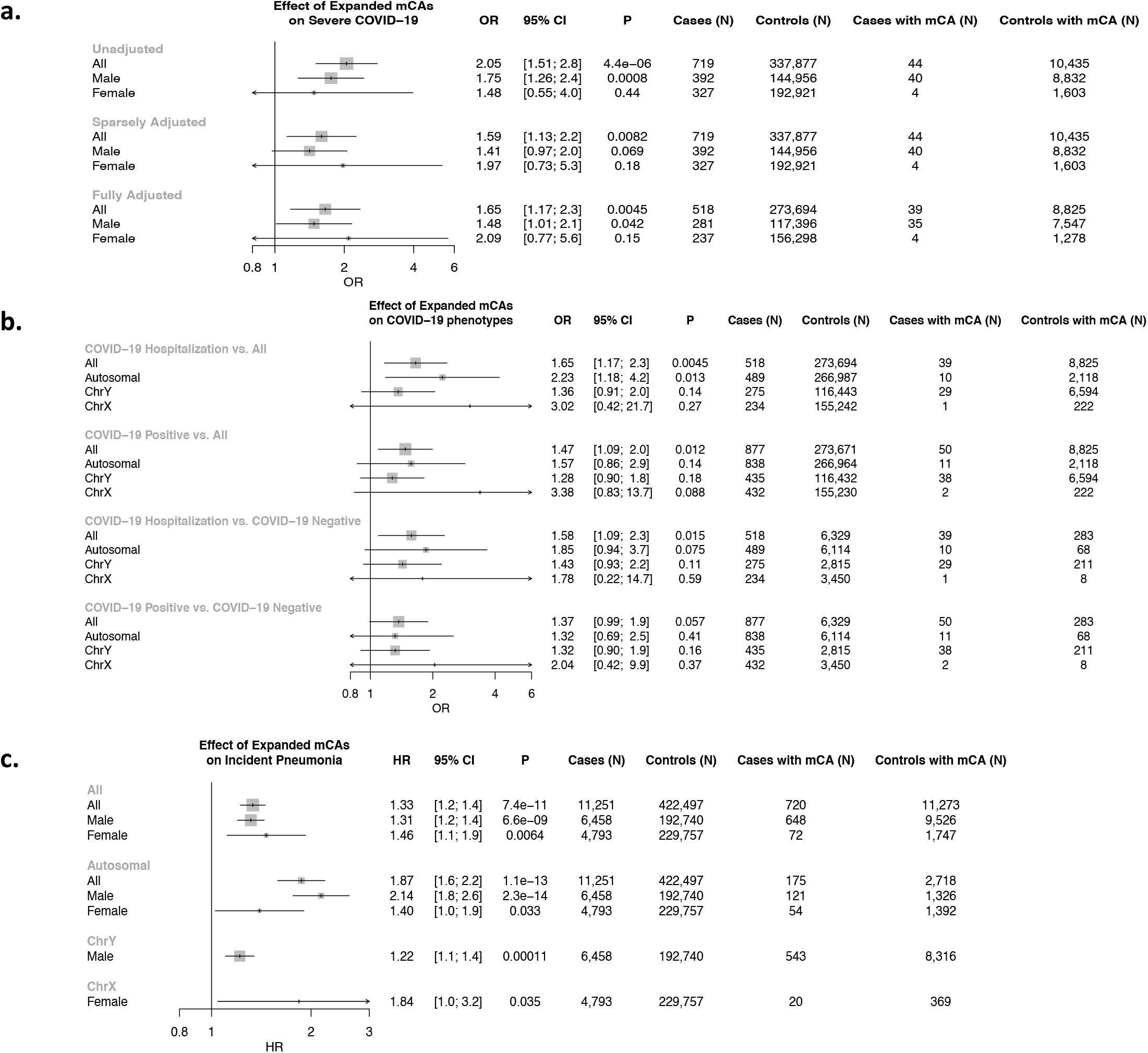
Associations of expanded mCAs in the UK Biobank with COVID-19 and incident pneumonia. Associations of expanded mCAs with **a**. COVID-19 hospitalization across different adjustment models, and **b**. different COVID-19 phenotypes in a fully adjusted model. Adjustment models include 1) an unadjusted model, 2) a sparsely adjusted model which adjusts for age, age2, sex, smoking status, and principal components of ancestry, and 3) a fully adjusted model which additionally adjusts for Townsend deprivation index, BMI, and the following comorbidities: Asthma, COPD, CAD, T2D, any cancer, and HTN. mCA = mosaic chromosomal alterations, COPD = chronic obstructive pulmonary disease, CAD = coronary artery disease, T2D = type 2 diabetes mellitus. **c**. Association of expanded mCAs with incident pneumonia stratified by sex, adjusted for age, age^2^, sex (in the All model only), smoking status, and principal components of ancestry. mCA = mosaic chromosomal alterations

**Extended Data Figure 9:**
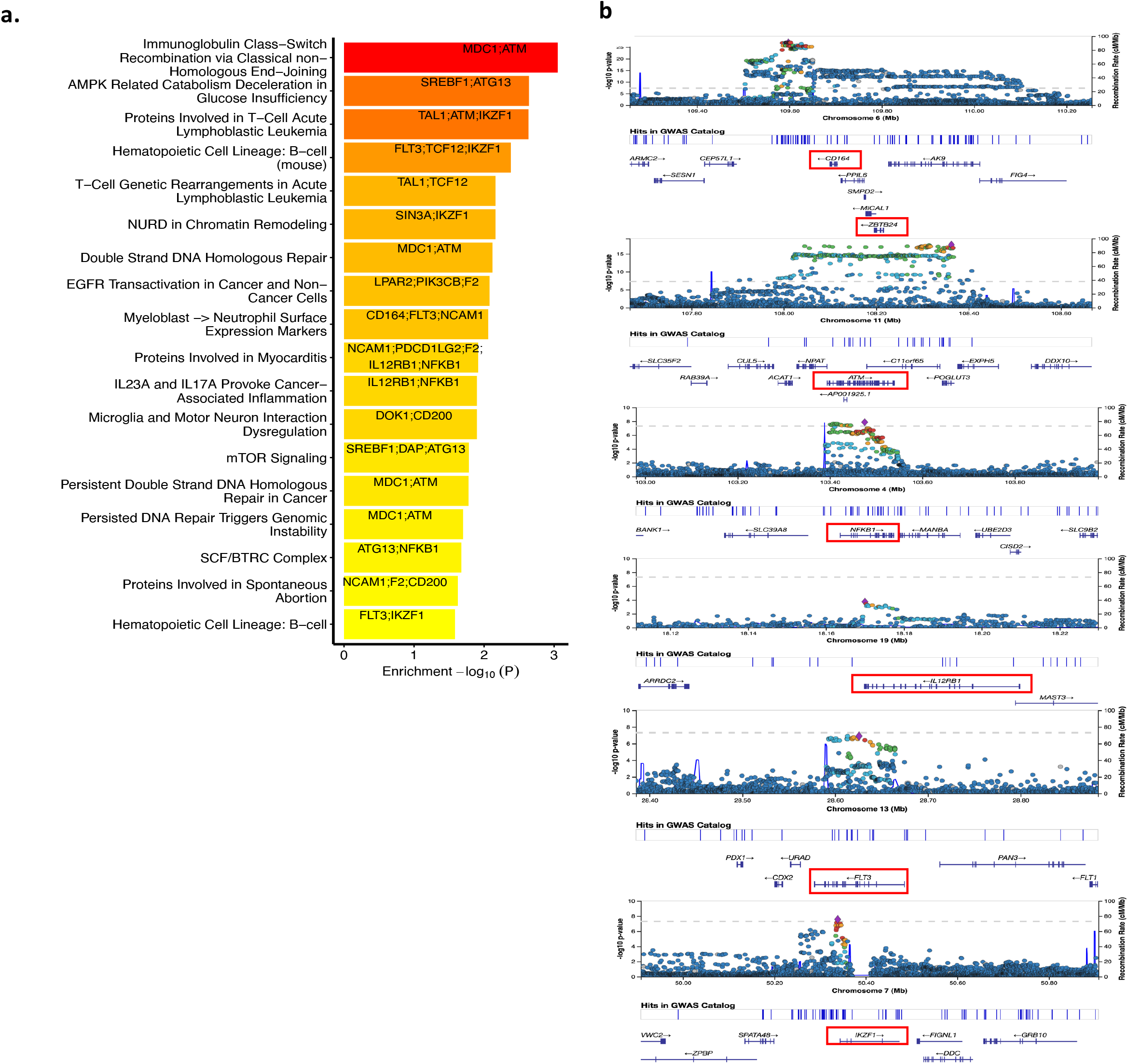
Pathway enrichment of TWAS results using the Elsevier Pathways. a. Top results from pathway enrichment analysis of the TWAS results using the Elsevier Pathways. b. Highlighting the GWAS locus-zoom plots for some of the TWAS genes implicated in the top pathways from panel b. Red boxes highlight the gene(s) with strongest association in the TWAS analyses. GWAS = genome-wide association study, TWAS = transcriptome-wide association study

**Extended Table 1:** Sensitivity analysis of expanded autosomal mCA with incident sepsis and with pneumonia association in the UK Biobank among those with cancer prior to incident infection, separately adjusting for chemotherapy, neutropenia, aplastic anemia, decreased white blood cell count, bone marrow or stem cell transplant, and radiation effects prior to infection (as defined using the Vanderbilt ICD-10 and ICD-9 phecode groupings^10^). Other covariates in the model included age, age^2^, sex, smoking status, and PC1-10 of ancestry. The summary stats (HR, P-value, 95% CI) reflect those for the expanded autosomal mCA term in each model. CI = confidence interval; HR = hazard ratio; mCA = mosaic chromosomal alteration; N = number; NOS = not otherwise specified

|  | *Adjustment* | *HR* | *P* | *Lower*<br>*95% CI* | *Upper*<br>*95% CI* |
| --- | --- | --- | --- | --- | --- |
| *Sepsis* | Chemotherapy | 2.48 | 3.04E-17 | 2.01 | 3.06 |
|  | Neutropenia | 1.65 | 3.98E-06 | 1.33 | 2.04 |
|  | Aplastic anemia | 2.58 | 1.84E-18 | 2.09 | 3.19 |
|  | Decreased white blood cell count | 1.65 | 3.98E-06 | 1.33 | 2.04 |
|  | Bone marrow or stem cell transplant | 2.77 | 3.25E-21 | 2.24 | 3.42 |
|  | Effects radiation NOS | 2.84 | 2.85E-22 | 2.30 | 3.51 |
| *Pneumonia* | Chemotherapy | 2.11 | 1.47E-14 | 1.74 | 2.55 |
|  | Neutropenia | 1.99 | 1.38E-12 | 1.65 | 2.41 |
|  | Aplastic anemia | 2.16 | 2.17E-15 | 1.79 | 2.62 |
|  | Decreased white blood cell count | 1.99 | 1.38E-12 | 1.65 | 2.41 |
|  | Bone marrow or stem cell transplant | 2.20 | 5.04E-16 | 1.82 | 2.66 |
|  | Effects radiation NOS | 2.27 | 2.59E-17 | 1.88 | 2.75 |

