## Supplementary Material for "Hematopoietic mosaic chromosomal alterations and risk for infection among 767,891 individuals without blood cancer"

**Table of Contents**

1) **Supplementary Note 1:** Extended Methods (pg.4-10)
   a) Cohorts
   b) Mosaic chromosomal alteration detection
   c) Other phenotype definitions
   d) Secondary statistical analyses
   e) Sensitivity analyses
2) Supplementary Figures (pg.11-26)
   a) **Supplementary Note 2:** MGB Biobank mCA sample quality control analyses.
   b) **Supplementary Note 3:** MGB Biobank mCA variant quality control analyses.
   c) **Supplementary Note 4**: FinnGen mCA sample quality control analyses.
   **d) Supplementary Note 5:** FinnGen mCA variant quality control analyses.
   e) **Supplementary Note 6:** Total number of mCAs (A) and expanded mCAs (B) per individual in the UK Biobank for mCA carriers.
   f) **Supplementary Note 7:** Prevalence of mCA categories by age bin across cohorts.
   g) **Supplementary Note 8:** Prevalence of expanded mCA categories by age bin across cohorts.
   h) **Supplementary Note 9**: Proportion of expanded autosomal mCAs carriers across lymphocyte count and percentage by age bin and sex.
   i) **Supplementary Note 10:** Associations of any mCA with incident infections.
   j) **Supplementary Note 11:** Suggestive associations (P<0.05) of expanded autosomal mCAs with incident infection categories.
   k) **Supplementary Note 12**: Associations of A) expanded ChrY and B) expanded ChrX mCAs with incident infections.
   l) **Supplementary Note 13:** Associations of expanded autosomal mCAs with incident sepsis and among different age strata in the UK Biobank.
   m) **Supplementary Note 14:** Associations of expanded autosomal mCAs with incident infections among A) those with antecedent cancer (ie: cancer prior to their infection) B) those without antecedent cancer.
   n) **Supplementary Note 15:** Interaction of expanded autosomal mCAs with antecedent cancer prior to infection in the UK Biobank and MGB Biobank.
   o) **Supplementary Note 16:** Correlated associations of 63 independent genome-wide significant variants associated with expanded mCAs (from Supplementary Data 3) between different mCA categories (expanded autosomal mCAs, expanded ChrX mCAs, expanded ChrY mCAs) in the UKB.
   p) **Supplementary Note 17:** Association of a mLOY PRS consisting of 156 previously identified[1] independent genome-wide significant variants associated with mLOY, with different expanded mCA categories in UKB dFemales.
3) Supplementary Tables (pg.27-33)
   a) **Supplementary Table 1:** Baseline summary statistics across the UK Biobank, MGB Biobank, and Biobank Japan among individuals analyzed.
   b) **Supplementary Table 2:** mCA counts by cohort.
   c) **Supplementary Table 3:** Association of potential risk factors with expanded autosomal mCAs in the UK Biobank.
   d) **Supplementary Table 4:** Association of mCAs with mortality from incident infection in Biobank Japan.

   e) **Supplementary Table 5:** Sensitivity analysis of incident sepsis association in the UK Biobank with the addition of a 25-factor smoking covariate, lymphocyte count and percentage, BMI, and prevalent type 2 diabetes.
   f) **Supplementary Table 6:** Sensitivity analysis of incident sepsis and pneumonia association in the UK Biobank among populations of individuals with different types of cancer prior to incident infection, where solid cancer is defined as any non-hematologic cancer.

4) **Supplementary Data:** (provided in external excel sheet)
   a) **Supplementary Data 1:** Infection phenotypic grouping categories across organ systems used in the UK Biobank, MGB Biobank, and Biobank Japan
   b) **Supplementary Data 2:** Infection phenotypic grouping categories across organ systems used in FinnGen (includes ICD10, 9, and 8 codes as specified).
   c) **Supplementary Data 3:** 63 independent loci identified in the expanded mCA GWAS
   d) **Supplementary Data 4:** Transcriptome-wide association using GTEXv8-whole blood (N=670)
   e) **Supplementary Data 5:** Gene set pathway enrichment analysis of the transcriptome-wide analyses (from Supplementary Data 4) using the Elsevier Pathway Collection through EnrichR
   f) **Supplementary Data 6:** Other phenotype definitions used in the UK Biobank COVID-19 sensitivity analyses
   g) **Supplementary Data 7:** Other phenotype definitions used in MGB Biobank
   h) **Supplementary Data 8:** Other phenotype definitions used in FinnGen

5) Supplementary References (pg. 34)

### Supplementary Note 1: Extended Methods

**Cohorts:**

The UK Biobank, a population-based cohort of approximately 500,000 participants recruited from 2006-2010, had existing genomic and longitudinal phenotypic data[2]. Baseline assessments were conducted at 22 assessment centres across the UK with sample collections including blood-derived DNA. Of 488,377 genotyped individuals, we analyzed 445,101 participants consenting to genetic analyses and who passed sample quality control criteria for mCA calling, had genotypic-phenotypic sex concordance, no 1$^{st}$ or 2$^{nd}$ degree relatives (random exclusion of one from each pair), and no prevalent hematologic cancer at time of blood draw. Genome-wide genotyping of blood-derived DNA was performed by UK Biobank using two genotyping arrays sharing 95% of marker content: Applied Biosystems UK BiLEVE Axiom Array (807,411 markers in 49,950 participants) and Applied Biosystems UK Biobank Axiom Array (825,927 markers in 438,427 participants) both by Affymetrix (Santa Clara, CA)[2]. Secondary use of the data was approved by the Massachusetts General Hospital Institutional Review Board (protocol 2013P001840) and facilitated through UK Biobank Applications 7089 and 21552.

The MGB Biobank (MGBB) contains genotypic and clinical data from >105,000 patients who consented to broad-based research across 7 regional hospitals[3]. Baseline phenotypes were ascertained from the electronic medical record (EMR) and surveys on lifestyle, environment, and family history. Of the approximately 36,000 genotyped individuals, 27,778 samples had available probe raw intensity data (IDAT) files for mCA calling. Blood-derived DNA samples were genotyped using three versions of the Multi-Ethnic Genotyping Array (MEGA) SNP array offered by Illumina. Secondary use of the data was approved by the Massachusetts General Hospital Institutional Review Board (protocol 2020P000904).

The FinnGen project (https://www.finngen.fi/en), launched in 2017, covers the whole of Finland and aims to improve health of people around the world through genetic studies. The latest released version (R6) contains genotypic, demographic, and extensive health (e.g. national inpatient/outpatient registers since 1969/1998, cancer register since 1953, and drug reimbursement register since 1964) information from 269,077 Finnish individuals. Blood-derived DNA samples were genotyped using two versions of FinnGen ThermoFisher Axiom custom array (https://www.finngen.fi/en/researchers/genotyping) provided by the Thermo Fisher genotyping service facility.

Biobank Japan (BBJ) is a hospital-based registry that collected clinical, DNA, and serum samples from approximately 200,000 consented patients with one or more of 47 target diseases at a total of 66 hospitals between 2003-2007[4]. Blood DNA was genotyped in three batches using different arrays or set of arrays, namely: (1) a combination of Illumina Infinium Omni Express and Human Exome; (2) Infinium Omni Express Exome v.1.0; and (3) Infinium Omni Express Exome v.1.2, which capture very similar SNPs. These analyses were approved by the ethics committees of RIKEN Center for Integrative Medical Sciences and the Institute of Medical Sciences, the University of Tokyo.

**Mosaic chromosomal alteration detection:**

Mosaic chromosomal alteration (mCA) detection in the UK Biobank was as described previously[5,6]. Briefly, genotype intensities were transformed to log2(R ratio) (LRR) and B-allele frequency (BAF values) to estimate total and relative allelic intensities, respectively. Re-phasing was performed using Eagle2[7] and mCA calling was performed by leveraging long-range phase information to search for allelic imbalances between maternal and paternal allelic fractions across contiguous genomic segments. Constitutional duplications and low-quality calls were filtered out and cell fraction was estimated as previously described[5]. UK Biobank mCA calls were obtained from dataset Return 2062 generated from UK Biobank application 19808.

Detection of mCAs in the MGB Biobank was performed starting from raw IDAT intensity files from the Illumina Multi-Ethnic Global Array (MEGA). Genotype clustering was performed using the Illumina GenCall algorithm. The resulting GTC genotype files were converted to VCF files using the bcftools gtc2vcf plugin (https://github.com/freeseek/gtc2vcf). Genotype phasing across the whole cohort was performed using SHAPEIT4[4] in windows of a maximum of 20 centimorgans with 2 centimorgans of overlap between consecutive windows. Phased genotypes were ligated across overlapping windows using bcftools concat (https://github.com/samtools/bcftools). mCA detection in the MGB Biobank was performed with MoChA[1,2] (https://github.com/freeseek/mocha). A pipeline to execute the whole workflow from raw files all the way to final mCA calls is available in WDL format for the Cromwell execution engine[8] as part of MoChA. We excluded 160 samples with phased B-allele frequency (BAF) auto-correlation >0.05, indicative of contamination or other potential sources of poor DNA quality, and 67 samples with phenotype-genotype sex discordance (**Supplementary Note 2**). We removed likely germline copy number polymorphisms (lod_baf_phase <20 for autosomal variants and lod_baf_phase <5 for sex chromosome variants), constitutional or inborn duplications (mCAs <2Mb with relative coverage >2.25, and mCAs 2-10Mb with relative coverage >2.4) and deletions (filtering out mCAs with relative coverage <0.5) (**Supplementary Note 3**).

FinnGen blood samples are genotyped by two versions of FinnGen ThermoFisher Axiom custom array. The detection of mCAs in FinnGen was performed starting from the genotype/intensity tables of 201,322 samples by using the "txt" mode of the MoChA WDL pipeline (https://github.com/freeseek/mocha). The input genotype/intensity tables for mCA detection were directly provided by the Thermo Fisher genotyping service, which performed genotype calling from the raw CEL files for each batch using the apt-probeset-genotype tool. Genotype phasing across the whole cohort was performed using SHAPEIT44 in windows of a maximum of 20 centimorgans with 2 centimorgans of overlap between consecutive windows. Phased genotypes were ligated across overlapping windows using bcftools concat (https://github.com/samtools/bcftools). We excluded 215 samples with phased B-allele frequency (BAF) auto-correlation >0.05, indicative of contamination or other potential sources of poor DNA quality, and 83 samples with phenotype-genotype sex discordance (**Supplementary Note 4**). We removed likely germline copy number polymorphisms (LOD_BAF_PHASE <20 for autosomal variants and LOD_BAF_PHASE <5 for sex chromosome variants, and LOD_BAF_PHASE <10 unless they are larger than 5Mbp (or 10 Mbp if they span the centromere)), constitutional or inborn duplications (0.5-5Mbp mCAs with relative coverage >2.5 and Bdev<0.1 and 5-10Mbp mCAs with relative coverage >2.75) and deletions (filtering out mCAs with relative coverage <0.5) (**Supplementary Note 5**). After further removing 1st or 2nd degree relatives, individuals with any prevalent hematologic cancer history at time of blood draw for genotyping, there were 175,690 samples remaining for analyses.

The detection of mCAs in the BBJ was as described previously[9]. Briefly, genotyping intensity data was analysed across variants shared between the three primary arrays, and used to compute BAF and LRR. Phasing was performed using the Eagle2 software. Mosaic events were called as previously described[5].

**Other phenotype definitions:**

Covariate definitions in the UK Biobank, including type 2 diabetes mellitus, hypertension, coronary artery disease, asthma, and chronic obstructive pulmonary disease, are provided in **Supplementary Data 6**. Cancer cases in the UK Biobank were identified using the cancer register (Category 100092) in combination with inpatient ICD-10 registry (Field IDs 41270/41280) and hematologic cancer cases were identified using the cancer registry's Field ID 40011 (hematological cancer identified from biopsy), Field ID 40005/40006 C81-96, D45-47, and inpatient ICD-10 registry (Field ID 41270/41280 C81-96, D45-47).

In the MGBB, cancer cases were identified using ICD-10 C00-D49, and hematologic cancer cases were identified using C81-96, D45-47. Other phenotype definitions in the MGBB are provided in **Supplementary Data 7**. Smoking status in MGBB was defined using a combination of electronic health record data and survey data. Follow-up time was coded as time from blood draw for genotyping to event (development of incident phenotype) or, for controls, time from sample collection to either censor date (10/31/19) or date of death if the patient died prior to the last censor.

Cancer and covariate phenotype definitions from FinnGen grouped across ICD 8, 9, and 10 codes are provided in **Supplementary Data 6**. Smoking status in FinGen was defined based on survey data. Follow-up time was coded as time from blood draw for genotyping to event (development of incident phenotype) or, for controls, time from sample collection to either censor date (12/31/19) or date of death if the patient died prior to the last censor.

**Secondary statistical analyses:**

Secondary associations were performed across other mCA exposures: all mCAs, all expanded autosomal mCAs, all autosomal mCAs, all ChrX mCAs, expanded ChrX mCAs, all ChrY mCAs, and expanded ChrY mCAs. Of note, approximately 99% of ChrX and ChrY mCA calls were loss of ChrX and ChrY (mLOX and mLOY, respectively). P-value threshold for significance among the secondary exposures was 0.05/10/7=0.0007. Additionally, associations of expanded autosomal mCAs with 20 secondary sub-outcomes separate from the organ system infections (listed under "category" in **Supplementary Data 1**) were performed to detect infection-specific associations. All association analyses were performed adjusted for age, $age^2$, sex, current or prior smoking history, and principal components 1-10 of genetic ancestry. P-value threshold for significance across these secondary analyses accounting for multiple hypothesis-testing was 0.05/20=0.0025.

**Sensitivity analyses:**

Further sensitivity analyses were performed in the UK Biobank expanded autosomal mCA and infection associations. First, stratified cancer analyses were performed among individuals with antecedent cancer prior to their incident infection in both the UK and MGB Biobanks, additionally stratifying for the same aforementioned covariates (age, age2, sex, ever smoking status, and the first ten principal components of genetic ancestry). Secondly, interaction analysis was performed using a mCA x antecedent cancer term in the model to analyze the interaction between mCAs and antecedent cancer prior to incident infection. Thirdly, for the incident sepsis association, adding four sets of covariates to the Cox proportional hazards model: 1) normalized body mass index and type 2 diabetes mellitus, 2) any antecedent cancer prior to incident infection, 3) adjusting for a more comprehensive 25-factor smoking phenotype[10], and 4) adjusting for normalized leukocyte count, lymphocyte count, and lymphocyte percentage at baseline visit. Fourthly, we evaluated the association of expanded autosomal mCAs incident sepsis and pneumonia associations among subgroups of individuals with cancer prior to infection including: prevalent solid cancer, incident hematologic cancer, and incident solid cancer prior to infection, in models adjusted for age, age2, sex, ever smoking status, and the first ten principal components of genetic ancestry. Lastly, we further evaluated the association of expanded autosomal mCAs with incident pneumonia and sepsis in separate models adjusted for different predictors of cancer morbidity including chemotherapy, neutropenia, aplastic anemia, decreased white blood cell count, bone marrow or stem cell transplant, and radiation effects prior to infection (with these phenotypes defined using the Vanderbilt ICD-10 and ICD-9 phecode groupings[11]), in the same aforementioned models adjusted for age, age$^2$, sex, ever smoking status, and the first ten principal components of genetic ancestry.

**Supplementary Figures**:

A.

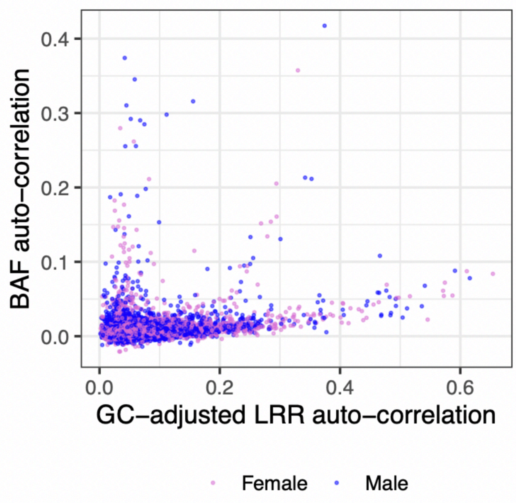


B.

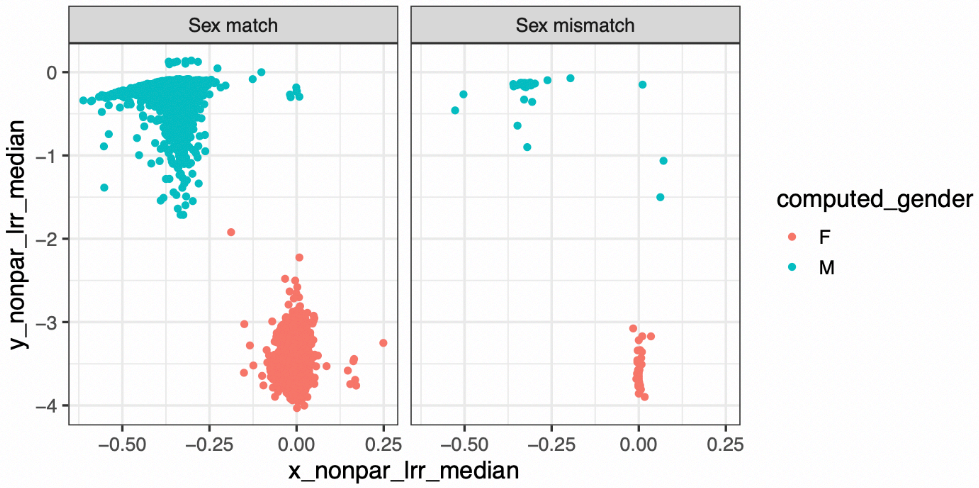


**Supplementary Note 2:** MGB Biobank mCA sample quality control analyses. A. plotting sample-level phased B allele frequency (BAF) auto-correlation across consecutive phased heterozygous sites versus Log R Ratio (LRR) of intensities using local GC content. B. Showing sex mismatches between MoChA-derived sex computed using the chrX nonPAR region versus reported sex. LRR=Log R Ratio, MoChA = mosaic chromosomal alterations caller, BAF=B allele frequency, nonPAR = Non-pseudoautosomal region

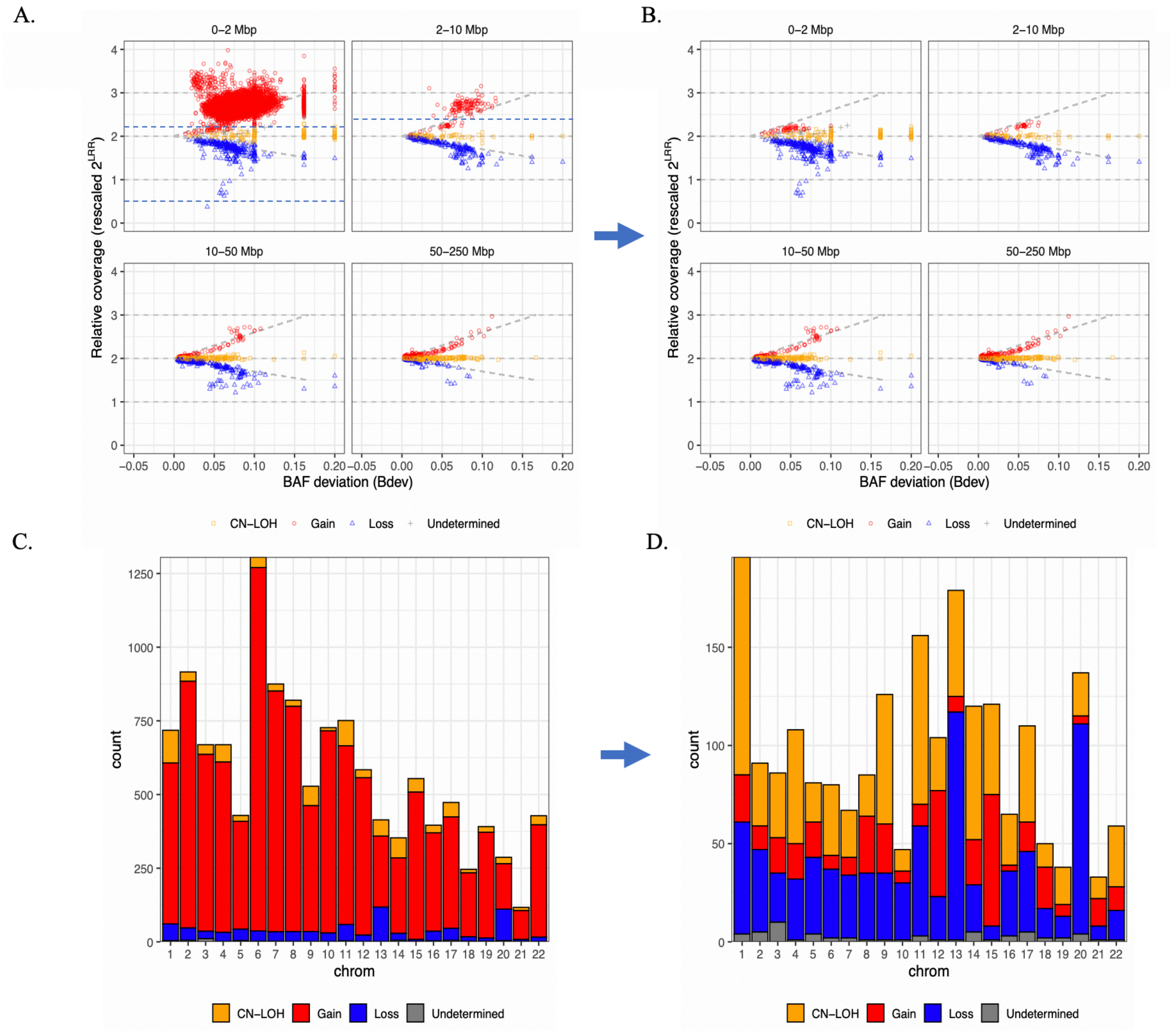


**Supplementary Note 3:** MGB Biobank mCA variant quality control analyses. Plots A. and C. represent mCAs carried among the quality-control filtered sample set, and after removal of likely germline variants (LOD_BAF_PHASE <20 for autosomal or mCAs annotated as known germline copy number polymorphisms). Plots B. and D. reflect additional variant quality control filters to remove constitutional duplications (0-2Mbp mCAs with relative coverage >2.25 and 2-10Mbp mCAs with relative coverage >2.4) and remove constitutional deletions (mCAs with relative coverage <0.5). CN-LOH = copy-neutral loss of heterozygosity, LOD_BAF_PHASE = log odds score for mCA model based on BAF and genotype phase, mCA = mosaic chromosomal alteration

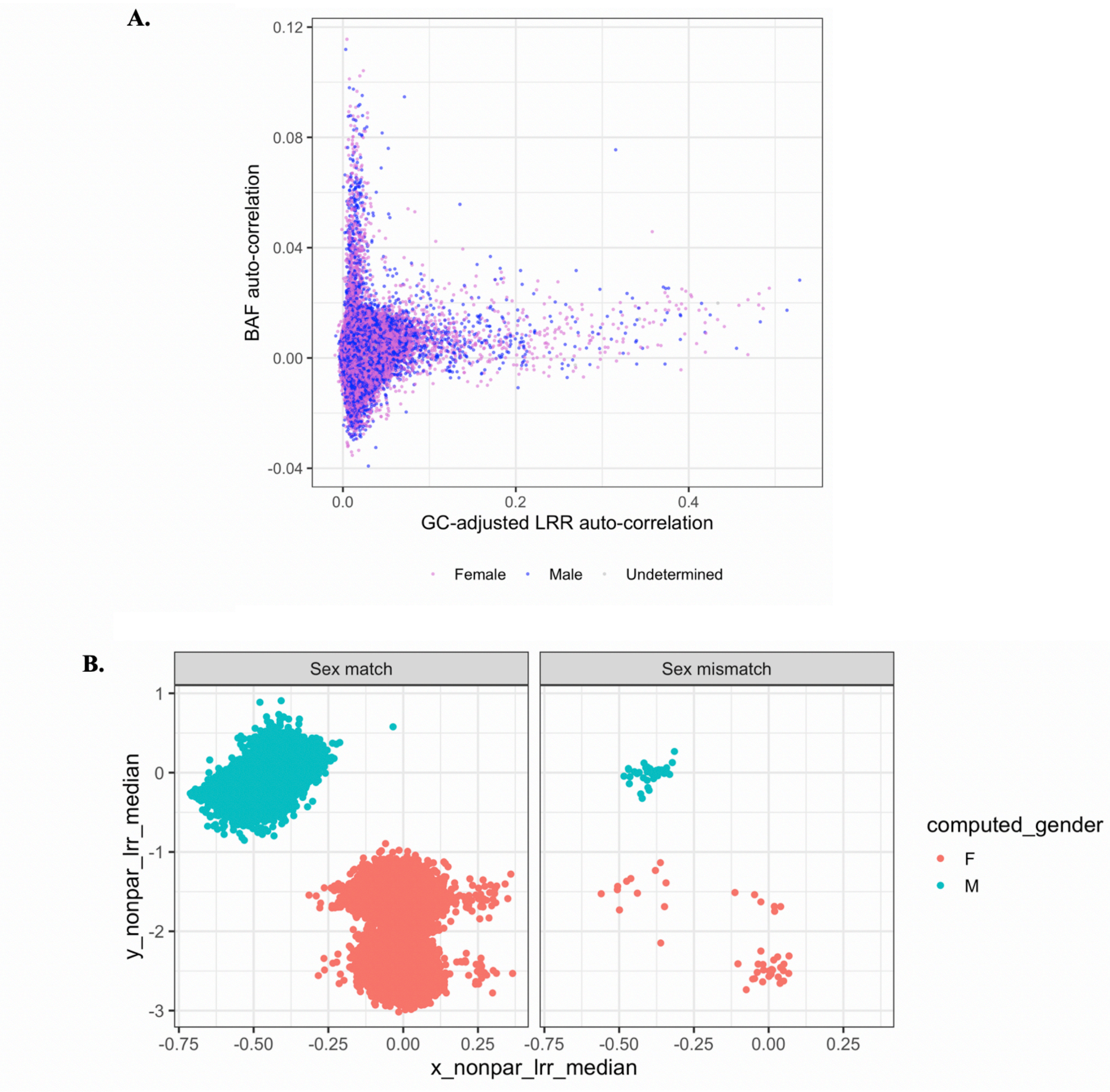


**Supplementary Note 4:** FinnGen mCA sample quality control analyses. A. plotting sample-level phased B allele frequency (BAF) auto-correlation across consecutive phased heterozygous sites versus Log R Ratio (LRR) of intensities using local GC content. B. Showing sex mismatches between MoChA-derived sex computed using the chrX nonPAR region versus reported sex. LRR=Log R Ratio, MoChA = mosaic chromosomal alterations caller, BAF=B allele frequency, nonPAR = Non-pseudoautosomal region

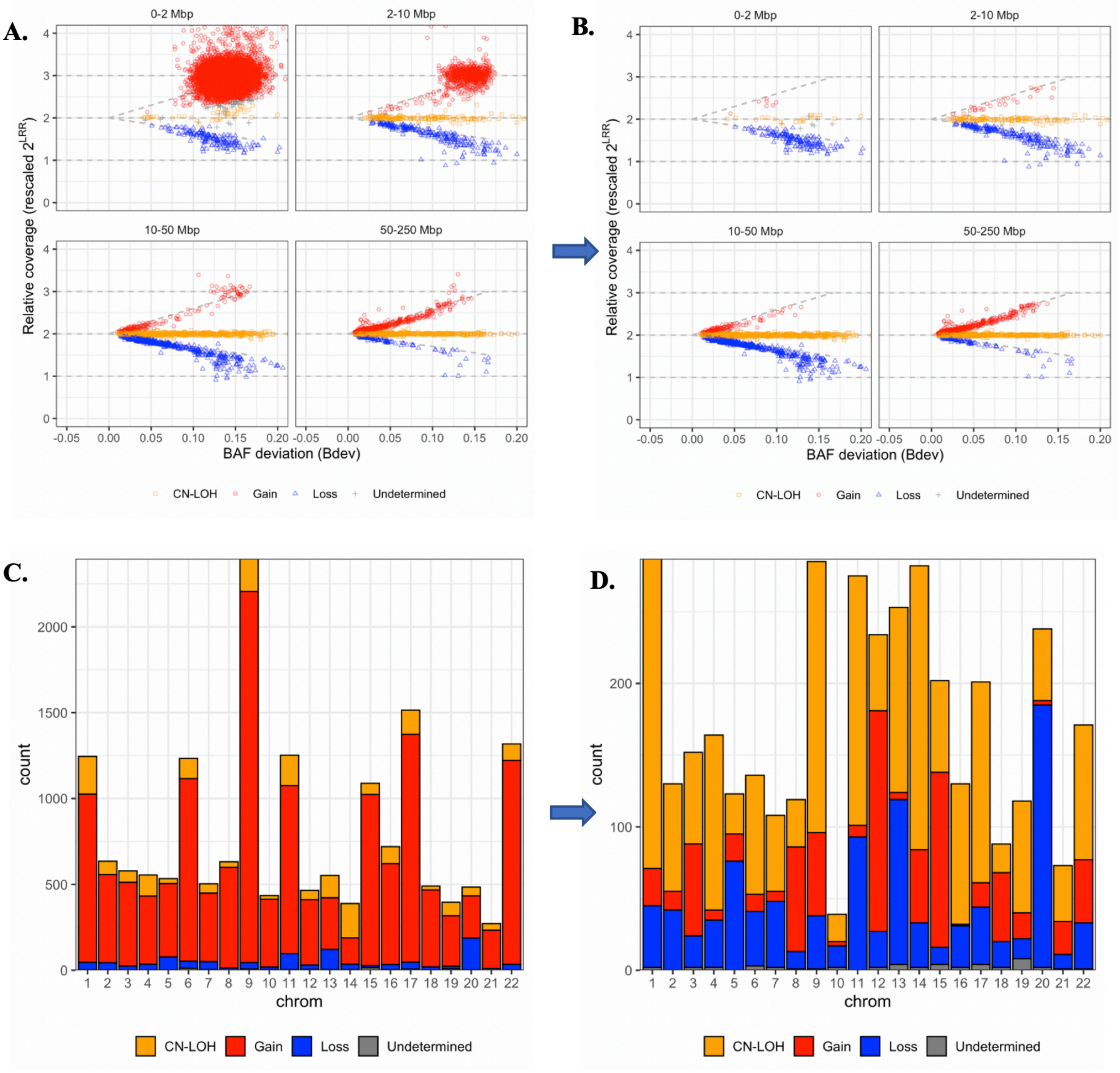


**Supplementary Note 5:** FinnGen mCA variant quality control analyses. Plots A. and C. represent mCAs after removal of likely germline variants (LOD_BAF_PHASE <20 for autosomal variants or mCAs annotated as known germline copy number polymorphisms). Plots B. and D. reflect additional sample quality control filters including removal of samples with call rate <0.97 and BAF_AUTO >0.03, and removal of calls with LOD_BAF_PHASE <10 unless they are larger than 5Mbp (or 10 Mbp if they span the centromere) and calls that are likely constitutional duplications (0.5-5Mbp mCAs with relative coverage >2.5 and Bdev<0.1 and 5-10Mbp mCAs with relative coverage >2.75) and constitutional deletions (mCAs with relative coverage <0.5). CN-LOH = copy-neutral loss of heterozygosity, LOD_BAF_PHASE = log odds score for mCA model based on BAF and genotype phase, mCA = mosaic chromosomal alteration

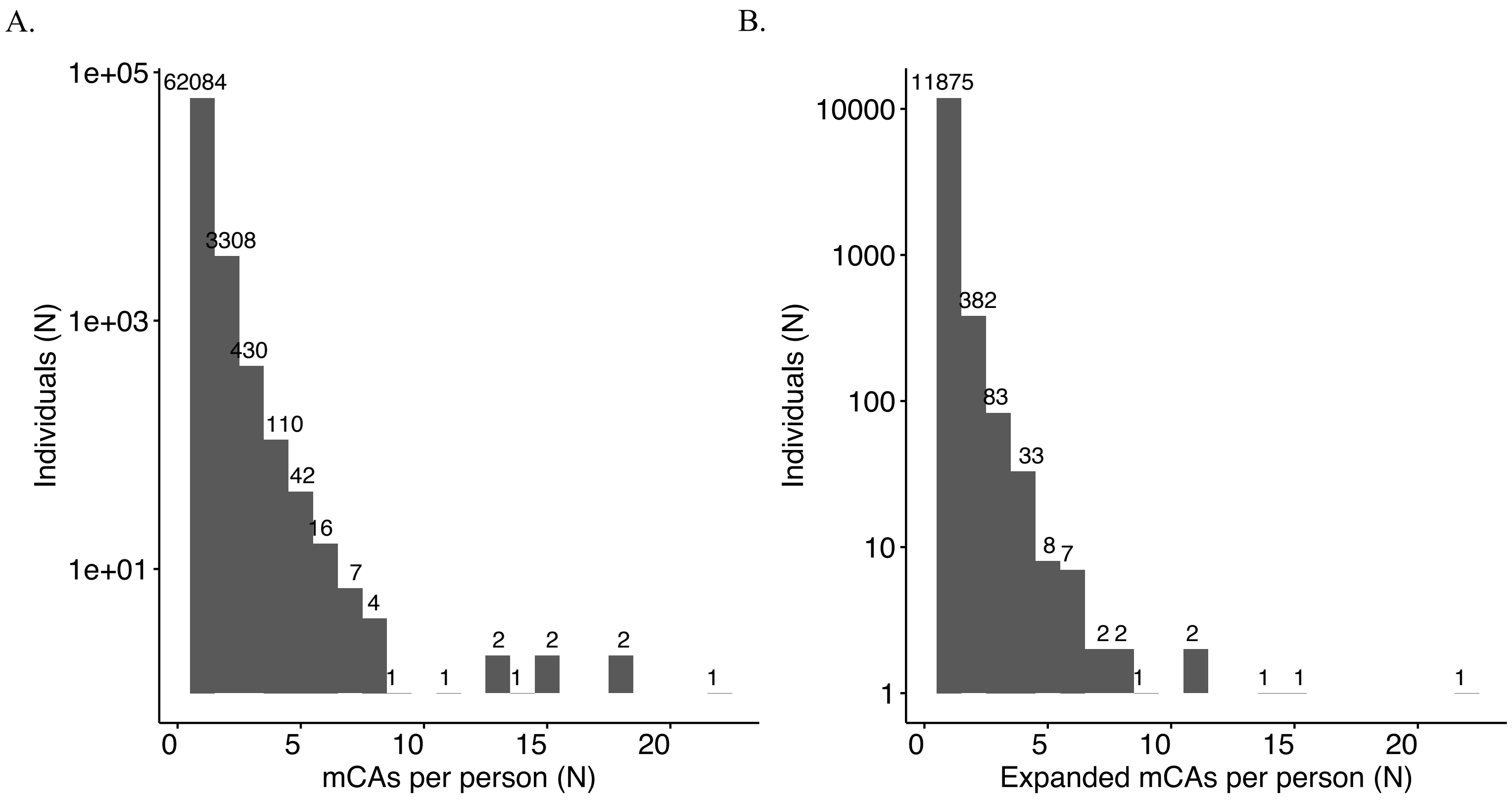


**Supplementary Note 6:** Total number of mCAs (A) and expanded mCAs (B) per individual in the UK Biobank for mCA carriers. mCAs = mosaic chromosomal alterations

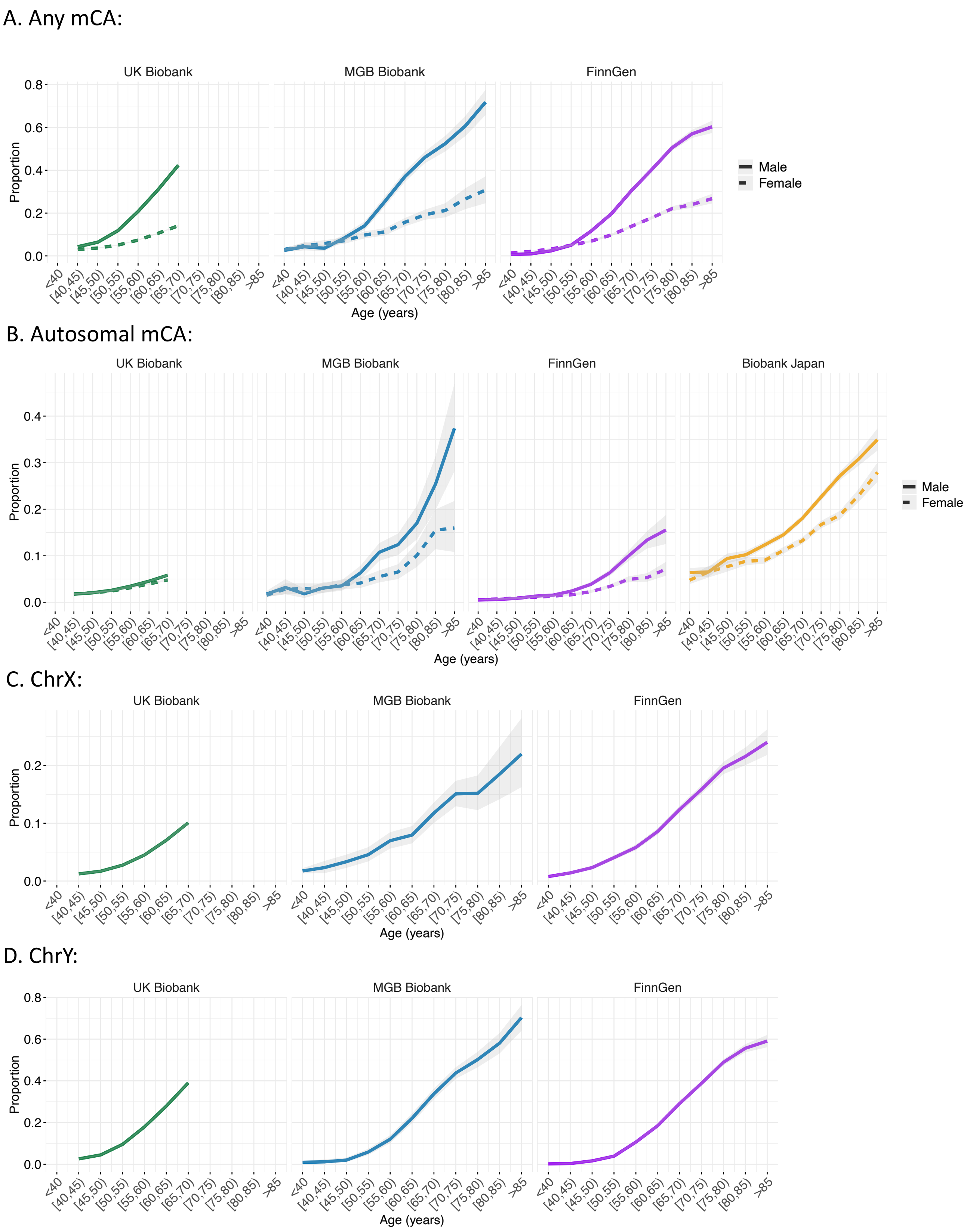


**Supplementary Note 7:** Prevalence of mCA categories by age bin across cohorts. mCA = mosaic chromosomal alterations.

A. Any expanded mCA:

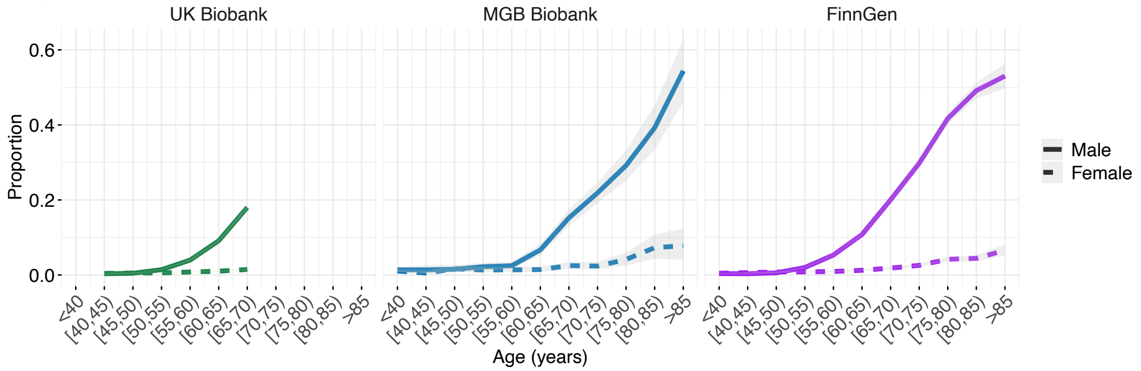


B. Expanded Autosomal mCA:

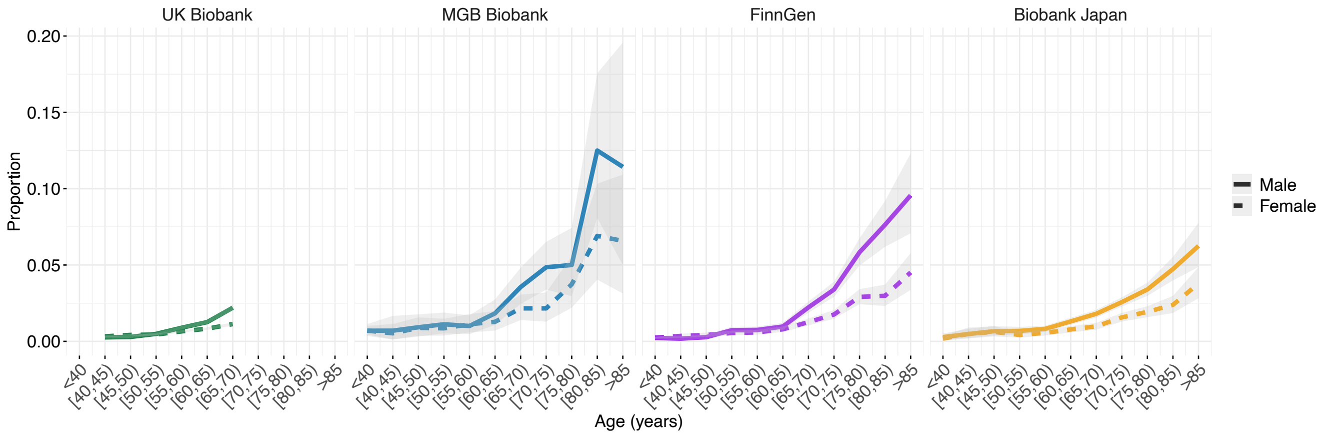


C. Expanded ChrX:

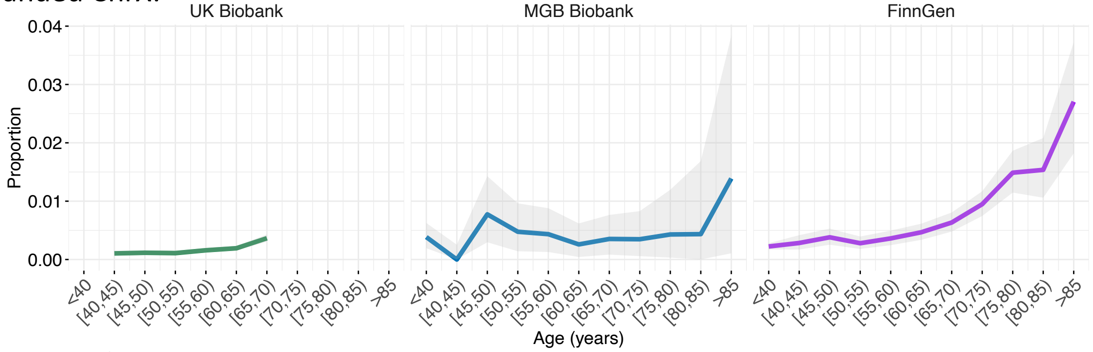


D. Expanded ChrY:

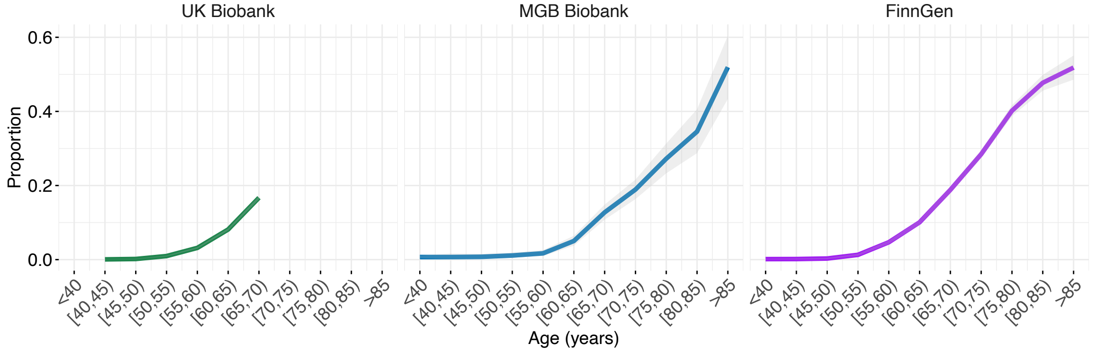


**Supplementary Note 8:** Prevalence of expanded mCA categories by age bin across cohorts. mCA = mosaic chromosomal alterations.

A.

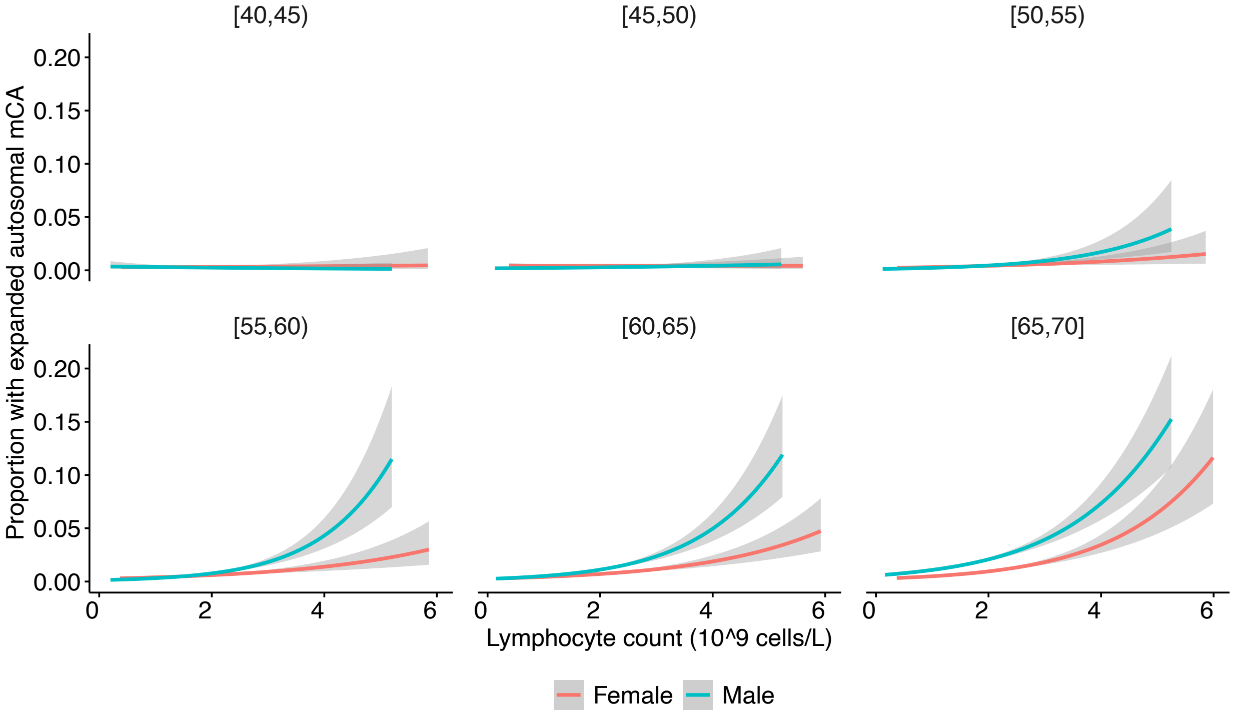


B.

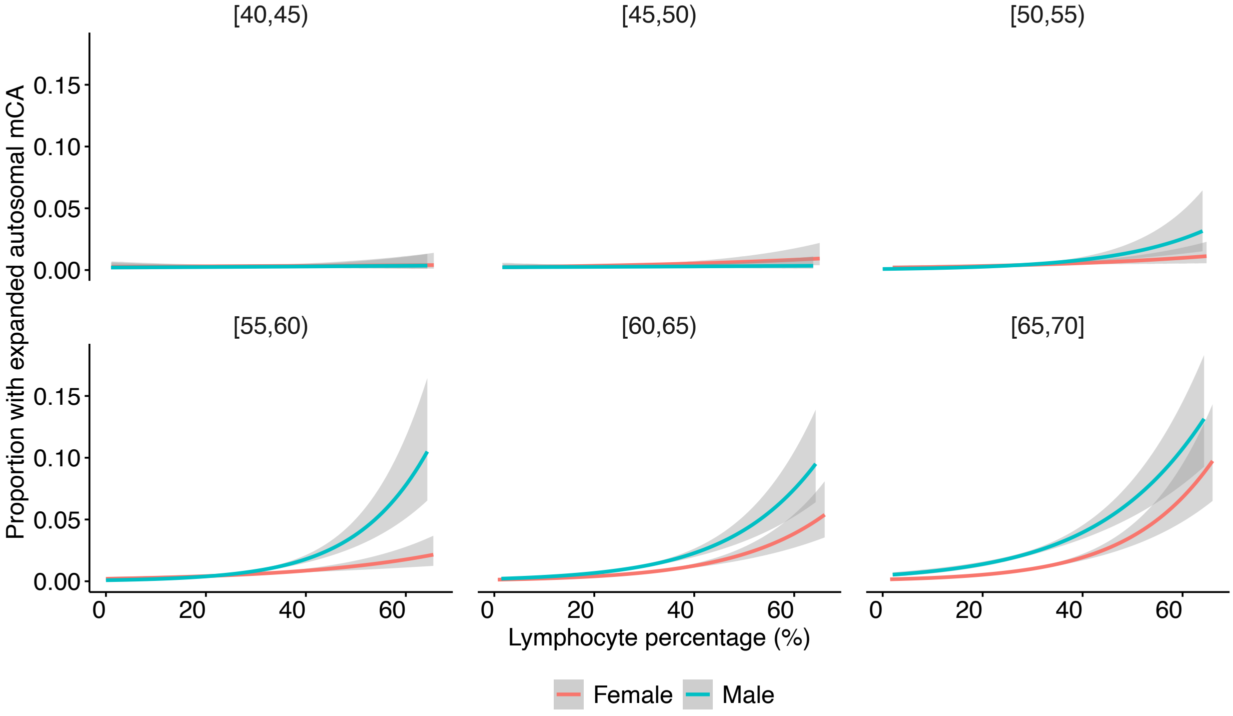


**Supplementary Note 9:** Proportion of expanded autosomal mCAs carriers across lymphocyte count and percentage by age bin and sex. Association of expanded autosomal mCA prevalence with A. Lymphocyte count and B. Lymphocyte percentage across bins of age and sex. mCA = mosaic chromosomal alterations.

A.

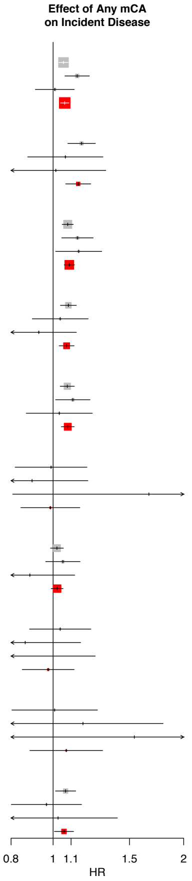


| | HR | 95% CI | P | Cases (N) | Controls (N) | Cases with mCA (N) | Controls with mCA (N) |
|---|---|---|---|---|---|---|---|
| **Any Infection** | | | | | | | |
| UK Biobank | 1.06 | [1.03; 1.08] | 1.3e−05 | 57,453 | 226,660 | 9,782 | 32,149 |
| MGB Biobank | 1.14 | [1.06; 1.21] | 0.00012 | 7,073 | 10,009 | 1,320 | 1,534 |
| FinnGen | 1.01 | [0.91; 1.12] | 0.86 | 3,608 | 30,810 | 462 | 3,251 |
| Overall | 1.06 | [1.04; 1.09] | 8.6e−08 | | | | |
| **Sepsis** | | | | | | | |
| UK Biobank | 1.16 | [1.08; 1.25] | 5.1e−05 | 5,237 | 437,757 | 1,185 | 64,592 |
| MGB Biobank | 1.07 | [0.87; 1.30] | 0.52 | 611 | 21,684 | 155 | 3,589 |
| FinnGen | 1.01 | [0.78; 1.32] | 0.92 | 448 | 64,860 | 75 | 6,765 |
| Overall | 1.14 | [1.07; 1.22] | 8.9e−05 | | | | |
| **Respiratory System Infection** | | | | | | | |
| UK Biobank | 1.08 | [1.05; 1.11] | 2.8e−07 | 39,158 | 320,202 | 6,843 | 46,417 |
| MGB Biobank | 1.14 | [1.05; 1.24] | 0.0024 | 4,411 | 16,249 | 798 | 2,707 |
| FinnGen | 1.15 | [1.01; 1.29] | 0.03 | 2,217 | 50,664 | 343 | 5,558 |
| Overall | 1.09 | [1.06; 1.12] | 5.9e−10 | | | | |
| **Digestive System Infection** | | | | | | | |
| UK Biobank | 1.08 | [1.04; 1.13] | 0.00017 | 19,681 | 403,978 | 3,418 | 59,280 |
| MGB Biobank | 1.04 | [0.90; 1.20] | 0.62 | 1,470 | 20,256 | 251 | 3,407 |
| FinnGen | 0.93 | [0.76; 1.13] | 0.46 | 1,091 | 58,323 | 122 | 6,406 |
| Overall | 1.07 | [1.03; 1.12] | 0.00042 | | | | |
| **Genitourinary System Infection** | | | | | | | |
| UK Biobank | 1.08 | [1.04; 1.12] | 9.2e−05 | 24,951 | 374,865 | 4,216 | 56,112 |
| MGB Biobank | 1.11 | [1.01; 1.22] | 0.025 | 3,617 | 16,859 | 658 | 2,781 |
| FinnGen | 1.03 | [0.87; 1.23] | 0.71 | 1,265 | 58,597 | 163 | 6,365 |
| Overall | 1.08 | [1.04; 1.12] | 8.4e−06 | | | | |
| **Cardiac Infection** | | | | | | | |
| UK Biobank | 0.99 | [0.82; 1.20] | 0.91 | 793 | 442,684 | 171 | 65,685 |
| MGB Biobank | 0.89 | [0.66; 1.20] | 0.46 | 280 | 22,031 | 69 | 3,670 |
| FinnGen | 1.66 | [0.81; 3.43] | 0.17 | 69 | 54,384 | 10 | 4,494 |
| Overall | 0.99 | [0.84; 1.15] | 0.85 | | | | |
| **Dermatologic Infection** | | | | | | | |
| UK Biobank | 1.02 | [0.99; 1.06] | 0.24 | 31,219 | 348,832 | 5,035 | 51,236 |
| MGB Biobank | 1.05 | [0.96; 1.15] | 0.27 | 3,588 | 17,319 | 671 | 2,827 |
| FinnGen | 0.88 | [0.70; 1.12] | 0.31 | 774 | 64,344 | 85 | 6,996 |
| Overall | 1.02 | [0.99; 1.05] | 0.18 | | | | |
| **Musculoskeletal System Infection** | | | | | | | |
| UK Biobank | 1.04 | [0.88; 1.22] | 0.65 | 1,307 | 441,037 | 241 | 65,462 |
| MGB Biobank | 0.86 | [0.64; 1.16] | 0.32 | 360 | 21,920 | 63 | 3,688 |
| FinnGen | 0.73 | [0.42; 1.25] | 0.25 | 152 | 68,041 | 16 | 7,391 |
| Overall | 0.97 | [0.85; 1.12] | 0.71 | | | | |
| **Nervous System Infection** | | | | | | | |
| UK Biobank | 1.01 | [0.80; 1.27] | 0.94 | 675 | 440,150 | 116 | 65,367 |
| MGB Biobank | 1.17 | [0.77; 1.79] | 0.47 | 168 | 22,210 | 30 | 3,740 |
| FinnGen | 1.54 | [0.75; 3.16] | 0.24 | 70 | 69,196 | 10 | 7,581 |
| Overall | 1.07 | [0.88; 1.30] | 0.48 | | | | |
| **Eye, Ear, or Mastoid Infection** | | | | | | | |
| UK Biobank | 1.07 | [1.01; 1.13] | 0.016 | 13,009 | 402,465 | 1,999 | 59,672 |
| MGB Biobank | 0.97 | [0.80; 1.16] | 0.71 | 1,008 | 21,134 | 151 | 3,575 |
| FinnGen | 1.03 | [0.75; 1.41] | 0.87 | 446 | 64,930 | 48 | 7,187 |
| Overall | 1.06 | [1.01; 1.11] | 0.026 | | | | |

**Supplementary Note 10:** Associations of A) any mCA and B) any expanded mCA with incident infections. mCA = mosaic chromosomal alterations.

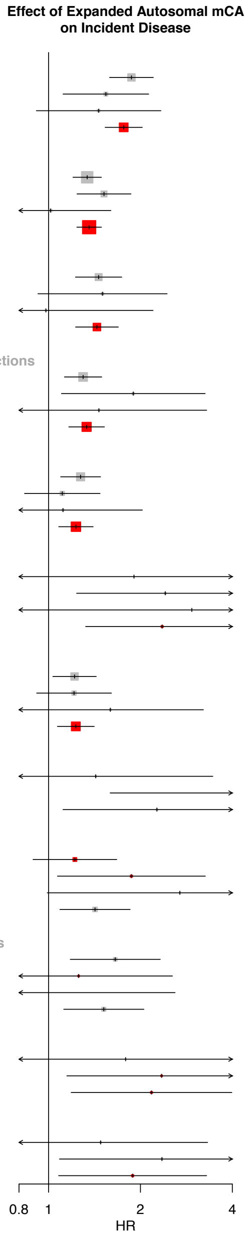


| | HR | 95% CI | P | Cases (N) | Controls (N) | Cases with mCA (N) | Controls with mCA (N) |
|---|---|---|---|---|---|---|---|
| **Pneumonia** | | | | | | | |
| UK Biobank | 1.87 | [1.58; 2.20] | 1.1e−13 | 11,251 | 422,497 | 175 | 2,718 |
| MGB Biobank | 1.54 | [1.12; 2.13] | 0.0087 | 1,516 | 20,409 | 39 | 286 |
| FinnGen | 1.46 | [0.91; 2.33] | 0.11 | 1,190 | 57,356 | 18 | 441 |
| Overall | 1.76 | [1.53; 2.03] | 2.3e−15 | | | | |
| **Any ICD A00–B99 Infection** | | | | | | | |
| UK Biobank | 1.34 | [1.20; 1.49] | 1e−07 | 42,452 | 324,344 | 408 | 2,078 |
| MGB Biobank | 1.52 | [1.24; 1.86] | 5.7e−05 | 5,047 | 14,842 | 97 | 192 |
| FinnGen | 1.02 | [0.64; 1.60] | 0.95 | 2,036 | 48,158 | 19 | 372 |
| Overall | 1.36 | [1.24; 1.49] | 1e−10 | | | | |
| **Gastroenteritis** | | | | | | | |
| UK Biobank | 1.46 | [1.22; 1.74] | 2.3e−05 | 14,621 | 420,331 | 149 | 2,768 |
| MGB Biobank | 1.50 | [0.92; 2.45] | 0.1 | 839 | 21,372 | 17 | 312 |
| FinnGen | 0.98 | [0.44; 2.20] | 0.96 | 620 | 57,281 | 6 | 473 |
| Overall | 1.44 | [1.23; 1.69] | 9e−06 | | | | |
| **Other Acute Lower Respiratory Infections** | | | | | | | |
| UK Biobank | 1.30 | [1.13; 1.49] | 0.00029 | 24,993 | 389,883 | 228 | 2,527 |
| MGB Biobank | 1.90 | [1.10; 3.26] | 0.021 | 489 | 21,823 | 14 | 317 |
| FinnGen | 1.46 | [0.65; 3.30] | 0.36 | 392 | 60,458 | 6 | 488 |
| Overall | 1.33 | [1.16; 1.52] | 2.8e−05 | | | | |
| **Pyelonephritis or UTI** | | | | | | | |
| UK Biobank | 1.27 | [1.10; 1.48] | 0.0017 | 21,929 | 388,914 | 210 | 2,533 |
| MGB Biobank | 1.11 | [0.83; 1.47] | 0.48 | 2,999 | 17,911 | 49 | 241 |
| FinnGen | 1.11 | [0.61; 2.03] | 0.72 | 862 | 58,456 | 11 | 470 |
| Overall | 1.23 | [1.08; 1.40] | 0.0018 | | | | |
| **Sexually Transmitted Infections** | | | | | | | |
| UK Biobank | 1.90 | [0.47; 7.70] | 0.37 | 248 | 441,724 | 3 | 2,971 |
| MGB Biobank | 2.41 | [1.24; 4.71] | 0.0099 | 378 | 21,900 | 9 | 327 |
| FinnGen | 2.95 | [0.40; 21.50] | 0.29 | 80 | 60,900 | 1 | 499 |
| Overall | 2.36 | [1.32; 4.20] | 0.0036 | | | | |
| **Acute Upper Respiratory Infections** | | | | | | | |
| UK Biobank | 1.22 | [1.03; 1.43] | 0.019 | 21,825 | 364,099 | 164 | 2,435 |
| MGB Biobank | 1.21 | [0.91; 1.61] | 0.18 | 3,353 | 17,891 | 50 | 269 |
| FinnGen | 1.59 | [0.79; 3.21] | 0.19 | 721 | 51,655 | 8 | 443 |
| Overall | 1.23 | [1.07; 1.41] | 0.0037 | | | | |
| **Anal Abscess** | | | | | | | |
| UK Biobank | 1.43 | [0.59; 3.45] | 0.43 | 730 | 441,579 | 6 | 2,964 |
| MGB Biobank | 5.22 | [1.59; 17.13] | 0.0064 | 62 | 22,382 | 3 | 333 |
| Overall | 2.26 | [1.11; 4.60] | 0.024 | | | | |
| **Conjunctivitis** | | | | | | | |
| UK Biobank | 1.22 | [0.89; 1.67] | 0.22 | 5,623 | 423,277 | 48 | 2,830 |
| MGB Biobank | 1.87 | [1.07; 3.26] | 0.028 | 556 | 21,769 | 13 | 321 |
| FinnGen | 2.69 | [0.99; 7.32] | 0.052 | 204 | 61,285 | 4 | 495 |
| Overall | 1.42 | [1.09; 1.85] | 0.0096 | | | | |
| **Appendicitis, peritonitis, pancreatitis** | | | | | | | |
| UK Biobank | 1.65 | [1.18; 2.32] | 0.0037 | 3,500 | 434,846 | 37 | 2,915 |
| MGB Biobank | 1.25 | [0.62; 2.55] | 0.53 | 453 | 21,792 | 8 | 325 |
| FinnGen | 0.36 | [0.05; 2.60] | 0.31 | 400 | 57,574 | 1 | 467 |
| Overall | 1.52 | [1.12; 2.05] | 0.0069 | | | | |
| **Hepatitis** | | | | | | | |
| UK Biobank | 1.79 | [0.57; 5.61] | 0.32 | 398 | 441,459 | 4 | 2,969 |
| MGB Biobank | 2.35 | [1.15; 4.79] | 0.019 | 305 | 21,867 | 8 | 325 |
| Overall | 2.17 | [1.19; 3.98] | 0.012 | | | | |
| **Osteomyelitis** | | | | | | | |
| UK Biobank | 1.48 | [0.66; 3.32] | 0.34 | 727 | 442,186 | 9 | 2,968 |
| MGB Biobank | 2.35 | [1.09; 5.10] | 0.03 | 183 | 22,197 | 7 | 329 |
| Overall | 1.89 | [1.08; 3.29] | 0.026 | | | | |

**Supplementary Note 11:** Suggestive associations (P<0.05) of expanded autosomal mCAs with incident infection categories. mCA = mosaic chromosomal alterations.

A.

**Effect of Expanded ChrX mCA on Incident Disease**

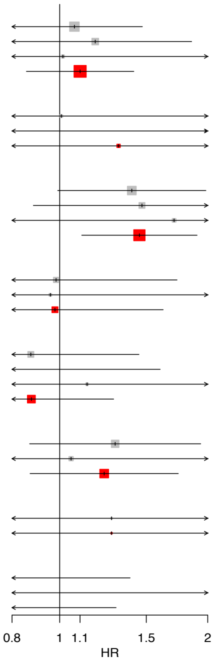


| | HR | 95% CI | P | Cases (N) | Controls (N) | Cases with mCA (N) | Controls with mCA (N) |
|---|---|---|---|---|---|---|---|
| **Any Infection** | | | | | | | |
| UK Biobank | 1.07 | [0.78; 1.5] | 0.67 | 29,681 | 120,124 | 68 | 191 |
| MGB Biobank | 1.18 | [0.75; 1.9] | 0.47 | 4,022 | 5,118 | 19 | 14 |
| FinnGen | 1.02 | [0.38; 2.7] | 0.98 | 975 | 28,393 | 4 | 109 |
| Overall | 1.10 | [0.86; 1.4] | 0.46 | | | | |
| **Sepsis** | | | | | | | |
| UK Biobank | 1.01 | [0.32; 3.1] | 0.99 | 2,392 | 236,677 | 4 | 393 |
| MGB Biobank | 1.98 | [0.49; 8.0] | 0.34 | 258 | 11,834 | 2 | 42 |
| Overall | 1.32 | [0.55; 3.2] | 0.54 | | | | |
| **Respiratory System Infection** | | | | | | | |
| UK Biobank | 1.40 | [0.99; 2.0] | 0.056 | 20,500 | 170,299 | 49 | 268 |
| MGB Biobank | 1.47 | [0.88; 2.4] | 0.14 | 2,613 | 8,516 | 15 | 23 |
| FinnGen | 1.71 | [0.76; 3.8] | 0.19 | 908 | 28,323 | 6 | 109 |
| Overall | 1.45 | [1.11; 1.9] | 0.0068 | | | | |
| **Digestive System Infection** | | | | | | | |
| UK Biobank | 0.98 | [0.56; 1.7] | 0.95 | 10,592 | 218,622 | 22 | 361 |
| MGB Biobank | 0.96 | [0.31; 3.0] | 0.94 | 801 | 11,000 | 3 | 38 |
| Overall | 0.98 | [0.59; 1.6] | 0.93 | | | | |
| **Genitourinary System Infection** | | | | | | | |
| UK Biobank | 0.87 | [0.53; 1.4] | 0.6 | 14,948 | 191,707 | 31 | 312 |
| MGB Biobank | 0.80 | [0.40; 1.6] | 0.53 | 2,453 | 8,354 | 8 | 30 |
| FinnGen | 1.14 | [0.36; 3.5] | 0.83 | 622 | 28,423 | 3 | 111 |
| Overall | 0.88 | [0.60; 1.3] | 0.5 | | | | |
| **Dermatologic Infection** | | | | | | | |
| UK Biobank | 1.30 | [0.87; 1.9] | 0.2 | 16,349 | 189,881 | 38 | 305 |
| MGB Biobank | 1.05 | [0.53; 2.1] | 0.88 | 1,912 | 9,462 | 8 | 30 |
| Overall | 1.23 | [0.87; 1.7] | 0.24 | | | | |
| **Musculoskeletal System Infection** | | | | | | | |
| MGB Biobank | 1.27 | [0.18; 9.2] | 0.81 | 175 | 11,893 | 1 | 43 |
| Overall | 1.27 | [0.18; 9.2] | 0.81 | | | | |
| **Eye, Ear, or Mastoid Infection** | | | | | | | |
| UK Biobank | 0.52 | [0.20; 1.4] | 0.19 | 7,367 | 215,374 | 7 | 354 |
| MGB Biobank | 0.73 | [0.18; 3.0] | 0.66 | 626 | 11,361 | 2 | 40 |
| Overall | 0.58 | [0.26; 1.3] | 0.19 | | | | |

B.

**Effect of Expanded ChrY mCA on Incident Disease**

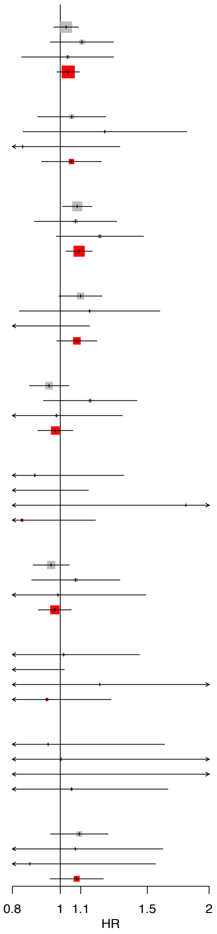


| | HR | 95% CI | P | Cases (N) | Controls (N) | Cases with mCA (N) | Controls with mCA (N) |
|---|---|---|---|---|---|---|---|
| **Any Infection** | | | | | | | |
| UK Biobank | 1.03 | [0.97; 1.1] | 0.36 | 27,772 | 106,536 | 1,506 | 4,425 |
| MGB Biobank | 1.11 | [0.95; 1.3] | 0.18 | 3,051 | 4,891 | 231 | 275 |
| FinnGen | 1.04 | [0.84; 1.3] | 0.75 | 1,156 | 21,300 | 110 | 1,798 |
| Overall | 1.04 | [0.98; 1.1] | 0.17 | | | | |
| **Sepsis** | | | | | | | |
| UK Biobank | 1.05 | [0.90; 1.2] | 0.51 | 2,845 | 201,080 | 201 | 8,929 |
| MGB Biobank | 1.23 | [0.84; 1.8] | 0.29 | 353 | 9,850 | 36 | 625 |
| FinnGen | 0.84 | [0.53; 1.3] | 0.45 | 244 | 25,875 | 25 | 2,119 |
| Overall | 1.05 | [0.92; 1.2] | 0.46 | | | | |
| **Respiratory System Infection** | | | | | | | |
| UK Biobank | 1.08 | [1.01; 1.2] | 0.024 | 18,658 | 149,903 | 1,093 | 6,374 |
| MGB Biobank | 1.07 | [0.89; 1.3] | 0.47 | 1,798 | 7,733 | 136 | 487 |
| FinnGen | 1.20 | [0.98; 1.5] | 0.075 | 1,096 | 18,599 | 124 | 1,707 |
| Overall | 1.09 | [1.03; 1.2] | 0.0051 | | | | |
| **Digestive System Infection** | | | | | | | |
| UK Biobank | 1.10 | [0.99; 1.2] | 0.067 | 9,089 | 185,356 | 508 | 8,154 |
| MGB Biobank | 1.15 | [0.83; 1.6] | 0.41 | 669 | 9,256 | 46 | 605 |
| FinnGen | 0.78 | [0.54; 1.1] | 0.21 | 513 | 23,145 | 33 | 2,059 |
| Overall | 1.08 | [0.98; 1.2] | 0.11 | | | | |
| **Genitourinary System Infection** | | | | | | | |
| UK Biobank | 0.95 | [0.87; 1.0] | 0.26 | 10,003 | 183,158 | 615 | 7,951 |
| MGB Biobank | 1.15 | [0.92; 1.4] | 0.21 | 1,164 | 8,505 | 110 | 502 |
| FinnGen | 0.98 | [0.72; 1.3] | 0.91 | 537 | 26,078 | 54 | 2,158 |
| Overall | 0.98 | [0.90; 1.1] | 0.58 | | | | |
| **Cardiac Infection** | | | | | | | |
| UK Biobank | 0.89 | [0.59; 1.3] | 0.57 | 487 | 203,599 | 33 | 9,112 |
| MGB Biobank | 0.57 | [0.29; 1.1] | 0.11 | 169 | 10,037 | 10 | 649 |
| FinnGen | 1.79 | [0.49; 6.6] | 0.38 | 37 | 21,162 | 3 | 1,269 |
| Overall | 0.84 | [0.59; 1.2] | 0.31 | | | | |
| **Dermatologic Infection** | | | | | | | |
| UK Biobank | 0.96 | [0.88; 1.0] | 0.32 | 14,870 | 158,951 | 711 | 7,178 |
| MGB Biobank | 1.07 | [0.88; 1.3] | 0.49 | 1,676 | 7,857 | 117 | 496 |
| FinnGen | 0.99 | [0.66; 1.5] | 0.96 | 390 | 25,670 | 29 | 2,223 |
| Overall | 0.97 | [0.90; 1.1] | 0.51 | | | | |
| **Musculoskeletal System Infection** | | | | | | | |
| UK Biobank | 1.02 | [0.71; 1.4] | 0.93 | 702 | 202,816 | 45 | 9,071 |
| MGB Biobank | 0.44 | [0.19; 1.0] | 0.055 | 185 | 10,027 | 6 | 661 |
| FinnGen | 1.20 | [0.57; 2.5] | 0.63 | 91 | 27,564 | 9 | 2,367 |
| Overall | 0.94 | [0.70; 1.3] | 0.68 | | | | |
| **Nervous System Infection** | | | | | | | |
| UK Biobank | 0.94 | [0.55; 1.6] | 0.84 | 338 | 202,604 | 18 | 9,081 |
| MGB Biobank | 1.00 | [0.38; 2.7] | 0.99 | 72 | 10,192 | 5 | 661 |
| FinnGen | 2.36 | [0.59; 9.4] | 0.22 | 27 | 28,056 | 3 | 2,418 |
| Overall | 1.05 | [0.67; 1.6] | 0.82 | | | | |
| **Eye, Ear, or Mastoid Infection** | | | | | | | |
| UK Biobank | 1.09 | [0.96; 1.2] | 0.2 | 5,642 | 187,091 | 272 | 8,297 |
| MGB Biobank | 1.07 | [0.71; 1.6] | 0.74 | 382 | 9,773 | 30 | 629 |
| FinnGen | 0.87 | [0.48; 1.6] | 0.63 | 199 | 26,390 | 14 | 2,303 |
| Overall | 1.08 | [0.95; 1.2] | 0.23 | | | | |

**Supplementary Note 15:** Associations of A) expanded ChrY and B) expanded ChrX mCAs with incident infections. mCA = mosaic chromosomal alterations.

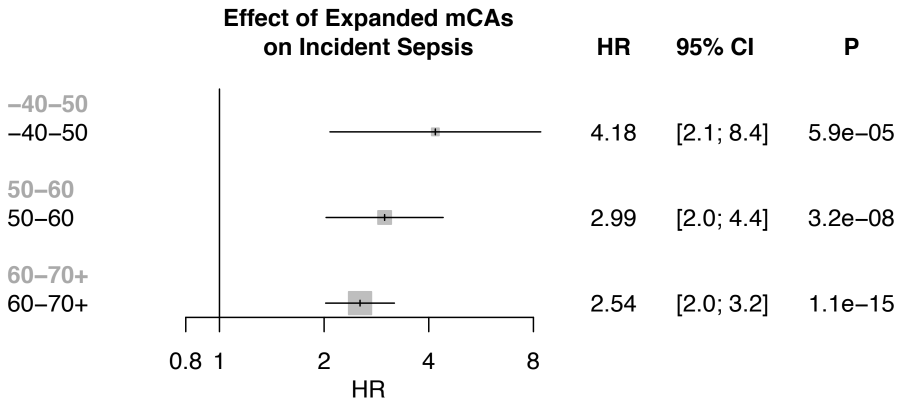


**Supplementary Note 13:** Associations of expanded autosomal mCAs with incident sepsis and among different age strata in the UK Biobank. Individuals with prevalent hematologic cancer were excluded from analyses. Associations were adjusted for sex, ever smoking status, and principal components 1-10 of ancestry. mCA = mosaic chromosomal alterations.

A.

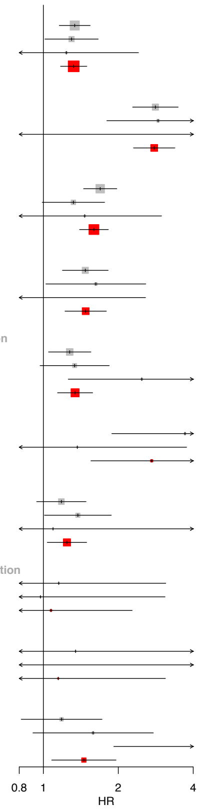


| +Prior Cancer History | HR | 95% CI | P | Cases (N) | Controls (N) | Cases with mCA (N) | Controls with mCA (N) |
|---|---|---|---|---|---|---|---|
| **Any Infection** | | | | | | | |
| UK Biobank | 1.34 | [1.16; 1.54] | 7e−05 | 17,180 | 53,991 | 231 | 471 |
| MGB Biobank | 1.30 | [1.01; 1.66] | 0.038 | 3,155 | 3,955 | 66 | 60 |
| FinnGen | 1.24 | [0.63; 2.41] | 0.53 | 550 | 3,158 | 9 | 48 |
| Overall | 1.32 | [1.17; 1.49] | 6.2e−06 | | | | |
| **Sepsis** | | | | | | | |
| UK Biobank | 2.82 | [2.28; 3.48] | 5.3e−22 | 3,575 | 119,106 | 99 | 1,131 |
| MGB Biobank | 2.89 | [1.80; 4.65] | 1.2e−05 | 383 | 11,338 | 19 | 185 |
| FinnGen | 0.57 | [0.08; 4.11] | 0.57 | 100 | 6,834 | 1 | 118 |
| Overall | 2.79 | [2.30; 3.38] | 9.7e−26 | | | | |
| **Respiratory System Infection** | | | | | | | |
| UK Biobank | 1.69 | [1.45; 1.97] | 2.1e−11 | 11,716 | 84,024 | 195 | 748 |
| MGB Biobank | 1.32 | [0.99; 1.76] | 0.059 | 2,296 | 7,825 | 48 | 134 |
| FinnGen | 1.47 | [0.72; 2.98] | 0.29 | 369 | 5,888 | 8 | 90 |
| Overall | 1.60 | [1.40; 1.82] | 6.1e−12 | | | | |
| **Digestive System Infection** | | | | | | | |
| UK Biobank | 1.47 | [1.19; 1.82] | 0.00032 | 7,064 | 108,613 | 107 | 1,048 |
| MGB Biobank | 1.62 | [1.02; 2.58] | 0.04 | 743 | 10,494 | 19 | 176 |
| FinnGen | 0.36 | [0.05; 2.57] | 0.31 | 166 | 6,595 | 1 | 116 |
| Overall | 1.48 | [1.22; 1.79] | 6.2e−05 | | | | |
| **Genitourinary System Infection** | | | | | | | |
| UK Biobank | 1.28 | [1.05; 1.55] | 0.015 | 9,159 | 96,744 | 127 | 953 |
| MGB Biobank | 1.34 | [0.97; 1.84] | 0.076 | 1,921 | 8,169 | 39 | 125 |
| FinnGen | 2.49 | [1.26; 4.91] | 0.0087 | 219 | 6,187 | 9 | 106 |
| Overall | 1.34 | [1.14; 1.58] | 0.0004 | | | | |
| **Cardiac Infection** | | | | | | | |
| UK Biobank | 3.71 | [1.88; 7.29] | 0.00015 | 271 | 122,774 | 9 | 1,225 |
| MGB Biobank | 1.37 | [0.50; 3.76] | 0.54 | 141 | 11,607 | 4 | 200 |
| Overall | 2.72 | [1.55; 4.78] | 0.00048 | | | | |
| **Dermatologic Infection** | | | | | | | |
| UK Biobank | 1.18 | [0.94; 1.48] | 0.15 | 7,668 | 94,777 | 94 | 911 |
| MGB Biobank | 1.38 | [1.01; 1.87] | 0.043 | 1,948 | 8,255 | 42 | 130 |
| FinnGen | 1.09 | [0.27; 4.49] | 0.9 | 138 | 7,160 | 2 | 117 |
| Overall | 1.24 | [1.04; 1.49] | 0.019 | | | | |
| **Musculoskeletal System Infection** | | | | | | | |
| UK Biobank | 1.15 | [0.43; 3.10] | 0.78 | 390 | 122,291 | 8 | 1,221 |
| MGB Biobank | 0.97 | [0.31; 3.08] | 0.96 | 167 | 11,589 | 3 | 199 |
| Overall | 1.07 | [0.51; 2.27] | 0.85 | | | | |
| **Nervous System Infection** | | | | | | | |
| UK Biobank | 1.35 | [0.43; 4.23] | 0.61 | 245 | 122,005 | 3 | 1,220 |
| MGB Biobank | 0.71 | [0.10; 5.15] | 0.73 | 88 | 11,740 | 1 | 206 |
| Overall | 1.15 | [0.43; 3.09] | 0.79 | | | | |
| **Eye, Ear, or Mastoid Infection** | | | | | | | |
| UK Biobank | 1.18 | [0.81; 1.72] | 0.38 | 2,866 | 110,982 | 33 | 1,117 |
| MGB Biobank | 1.58 | [0.91; 2.77] | 0.11 | 543 | 10,992 | 13 | 193 |
| FinnGen | 5.50 | [1.92; 15.76] | 0.0015 | 58 | 7,519 | 4 | 123 |
| Overall | 1.45 | [1.08; 1.96] | 0.014 | | | | |

B.

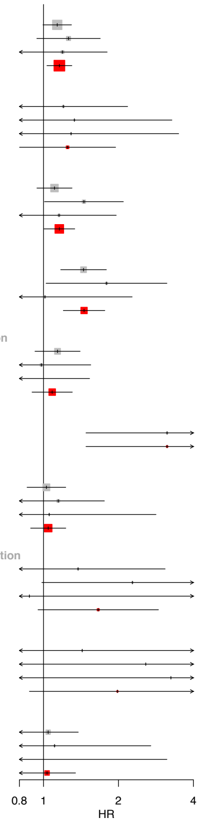


| −Prior Cancer History | HR | 95% CI | P | Cases (N) | Controls (N) | Cases with mCA (N) | Controls with mCA (N) |
|---|---|---|---|---|---|---|---|
| **Any Infection** | | | | | | | |
| UK Biobank | 1.14 | [0.99; 1.30] | 0.06 | 40,273 | 172,669 | 266 | 921 |
| MGB Biobank | 1.26 | [0.94; 1.69] | 0.12 | 3,918 | 6,054 | 46 | 64 |
| FinnGen | 1.19 | [0.79; 1.80] | 0.4 | 2,628 | 24,604 | 23 | 155 |
| Overall | 1.16 | [1.03; 1.30] | 0.013 | | | | |
| **Sepsis** | | | | | | | |
| UK Biobank | 1.20 | [0.66; 2.18] | 0.55 | 1,662 | 318,651 | 12 | 1,739 |
| MGB Biobank | 1.33 | [0.54; 3.28] | 0.54 | 228 | 10,346 | 5 | 123 |
| FinnGen | 1.29 | [0.48; 3.49] | 0.62 | 278 | 51,714 | 4 | 335 |
| Overall | 1.25 | [0.80; 1.95] | 0.33 | | | | |
| **Respiratory System Infection** | | | | | | | |
| UK Biobank | 1.11 | [0.94; 1.30] | 0.22 | 27,442 | 236,178 | 171 | 1,263 |
| MGB Biobank | 1.45 | [1.01; 2.09] | 0.045 | 2,115 | 8,424 | 30 | 94 |
| FinnGen | 1.15 | [0.68; 1.96] | 0.59 | 1,527 | 39,575 | 14 | 267 |
| Overall | 1.16 | [1.00; 1.34] | 0.045 | | | | |
| **Digestive System Infection** | | | | | | | |
| UK Biobank | 1.45 | [1.17; 1.79] | 0.0006 | 12,617 | 295,365 | 98 | 1,603 |
| MGB Biobank | 1.79 | [1.02; 3.13] | 0.041 | 727 | 9,762 | 13 | 115 |
| FinnGen | 1.01 | [0.45; 2.27] | 0.97 | 810 | 45,753 | 6 | 315 |
| Overall | 1.46 | [1.20; 1.76] | 0.00013 | | | | |
| **Genitourinary System Infection** | | | | | | | |
| UK Biobank | 1.14 | [0.92; 1.40] | 0.22 | 15,792 | 278,121 | 105 | 1,489 |
| MGB Biobank | 0.98 | [0.62; 1.55] | 0.93 | 1,696 | 8,690 | 19 | 103 |
| FinnGen | 0.57 | [0.21; 1.53] | 0.27 | 896 | 46,468 | 4 | 317 |
| Overall | 1.08 | [0.90; 1.31] | 0.4 | | | | |
| **Cardiac Infection** | | | | | | | |
| MGB Biobank | 3.14 | [1.48; 6.64] | 0.0028 | 139 | 10,424 | 8 | 121 |
| Overall | 3.14 | [1.48; 6.64] | 0.0028 | | | | |
| **Dermatologic Infection** | | | | | | | |
| UK Biobank | 1.03 | [0.86; 1.23] | 0.77 | 23,551 | 254,055 | 145 | 1,411 |
| MGB Biobank | 1.15 | [0.75; 1.76] | 0.53 | 1,640 | 9,064 | 22 | 112 |
| FinnGen | 1.05 | [0.39; 2.83] | 0.92 | 557 | 50,648 | 4 | 343 |
| Overall | 1.04 | [0.89; 1.23] | 0.6 | | | | |
| **Musculoskeletal System Infection** | | | | | | | |
| UK Biobank | 1.38 | [0.62; 3.08] | 0.44 | 917 | 318,746 | 8 | 1,736 |
| MGB Biobank | 2.28 | [0.98; 5.26] | 0.054 | 193 | 10,331 | 6 | 126 |
| FinnGen | 0.88 | [0.12; 6.33] | 0.9 | 124 | 53,436 | 1 | 367 |
| Overall | 1.66 | [0.95; 2.90] | 0.075 | | | | |
| **Nervous System Infection** | | | | | | | |
| UK Biobank | 1.43 | [0.46; 4.47] | 0.54 | 430 | 318,145 | 3 | 1,738 |
| MGB Biobank | 2.57 | [0.61; 10.81] | 0.2 | 80 | 10,470 | 2 | 126 |
| FinnGen | 3.25 | [0.44; 23.94] | 0.25 | 54 | 54,233 | 1 | 370 |
| Overall | 1.98 | [0.88; 4.47] | 0.1 | | | | |
| **Eye, Ear, or Mastoid Infection** | | | | | | | |
| UK Biobank | 1.04 | [0.79; 1.38] | 0.76 | 10,143 | 291,483 | 59 | 1,596 |
| MGB Biobank | 1.11 | [0.45; 2.70] | 0.82 | 465 | 10,142 | 5 | 123 |
| FinnGen | 0.44 | [0.06; 3.13] | 0.41 | 345 | 50,706 | 1 | 359 |
| Overall | 1.03 | [0.79; 1.34] | 0.81 | | | | |

**Supplementary Note 14:** Associations of expanded autosomal mCAs with incident infections among A) those with antecedent cancer (ie: cancer prior to their infection) B) those without antecedent cancer.  mCA = mosaic chromosomal alterations.

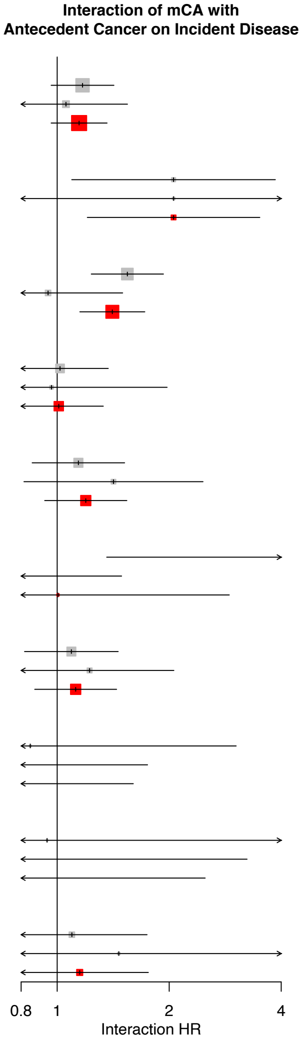


| | Interaction HR | Interaction 95% CI | Interaction P | Cases (N) | Controls (N) | Cases with mCA (N) | Controls with mCA (N) |
|---|---|---|---|---|---|---|---|
| **Any Infection** | | | | | | | |
| UK Biobank | 1.17 | [0.96; 1.42] | 0.11 | 57453 | 226660 | 497 | 1392 |
| MGB Biobank | 1.06 | [0.72; 1.54] | 0.78 | 7073 | 10009 | 112 | 124 |
| Overall | 1.15 | [0.96; 1.36] | 0.12 | | | | |
| **Sepsis** | | | | | | | |
| UK Biobank | 2.05 | [1.09; 3.86] | 0.025 | 5237 | 437757 | 111 | 2870 |
| MGB Biobank | 2.06 | [0.75; 5.61] | 0.16 | 611 | 21684 | 24 | 308 |
| Overall | 2.05 | [1.20; 3.50] | 0.0082 | | | | |
| **Respiratory System Infection** | | | | | | | |
| UK Biobank | 1.54 | [1.23; 1.93] | 0.00014 | 39158 | 320202 | 366 | 2011 |
| MGB Biobank | 0.95 | [0.60; 1.50] | 0.81 | 4411 | 16249 | 78 | 228 |
| Overall | 1.41 | [1.15; 1.72] | 0.00088 | | | | |
| **Digestive System Infection** | | | | | | | |
| UK Biobank | 1.02 | [0.76; 1.37] | 0.91 | 19681 | 403978 | 205 | 2651 |
| MGB Biobank | 0.97 | [0.47; 1.97] | 0.92 | 1470 | 20256 | 32 | 291 |
| Overall | 1.01 | [0.77; 1.33] | 0.94 | | | | |
| **Genitourinary System Infection** | | | | | | | |
| UK Biobank | 1.14 | [0.86; 1.52] | 0.37 | 24951 | 374865 | 232 | 2442 |
| MGB Biobank | 1.42 | [0.81; 2.47] | 0.22 | 3617 | 16859 | 58 | 228 |
| Overall | 1.19 | [0.93; 1.54] | 0.17 | | | | |
| **Cardiac Infection** | | | | | | | |
| UK Biobank | 10.82 | [1.36; 86.10] | 0.024 | 793 | 442684 | 10 | 2973 |
| MGB Biobank | 0.43 | [0.13; 1.49] | 0.18 | 280 | 22031 | 12 | 321 |
| Overall | 1.00 | [0.35; 2.90] | 0.99 | | | | |
| **Dermatologic Infection** | | | | | | | |
| UK Biobank | 1.09 | [0.82; 1.46] | 0.55 | 31219 | 348832 | 239 | 2322 |
| MGB Biobank | 1.22 | [0.73; 2.06] | 0.45 | 3588 | 17319 | 64 | 242 |
| Overall | 1.12 | [0.87; 1.44] | 0.38 | | | | |
| **Musculoskeletal System Infection** | | | | | | | |
| UK Biobank | 0.85 | [0.24; 3.03] | 0.8 | 1307 | 441037 | 16 | 2957 |
| MGB Biobank | 0.43 | [0.11; 1.75] | 0.24 | 360 | 21920 | 9 | 325 |
| Overall | 0.62 | [0.24; 1.60] | 0.33 | | | | |
| **Nervous System Infection** | | | | | | | |
| UK Biobank | 0.94 | [0.19; 4.71] | 0.94 | 675 | 440150 | 6 | 2958 |
| MGB Biobank | 0.29 | [0.03; 3.24] | 0.31 | 168 | 22210 | 3 | 332 |
| Overall | 0.65 | [0.17; 2.50] | 0.53 | | | | |
| **Eye, Ear, or Mastoid Infection** | | | | | | | |
| UK Biobank | 1.10 | [0.69; 1.74] | 0.7 | 13009 | 402465 | 92 | 2713 |
| MGB Biobank | 1.46 | [0.52; 4.15] | 0.47 | 1008 | 21134 | 18 | 316 |
| Overall | 1.15 | [0.75; 1.76] | 0.52 | | | | |

**Supplementary Note 15:** Interaction of expanded autosomal mCAs with antecedent cancer prior to infection in the UK Biobank and MGB Biobank. mCA = mosaic chromosomal alterations.

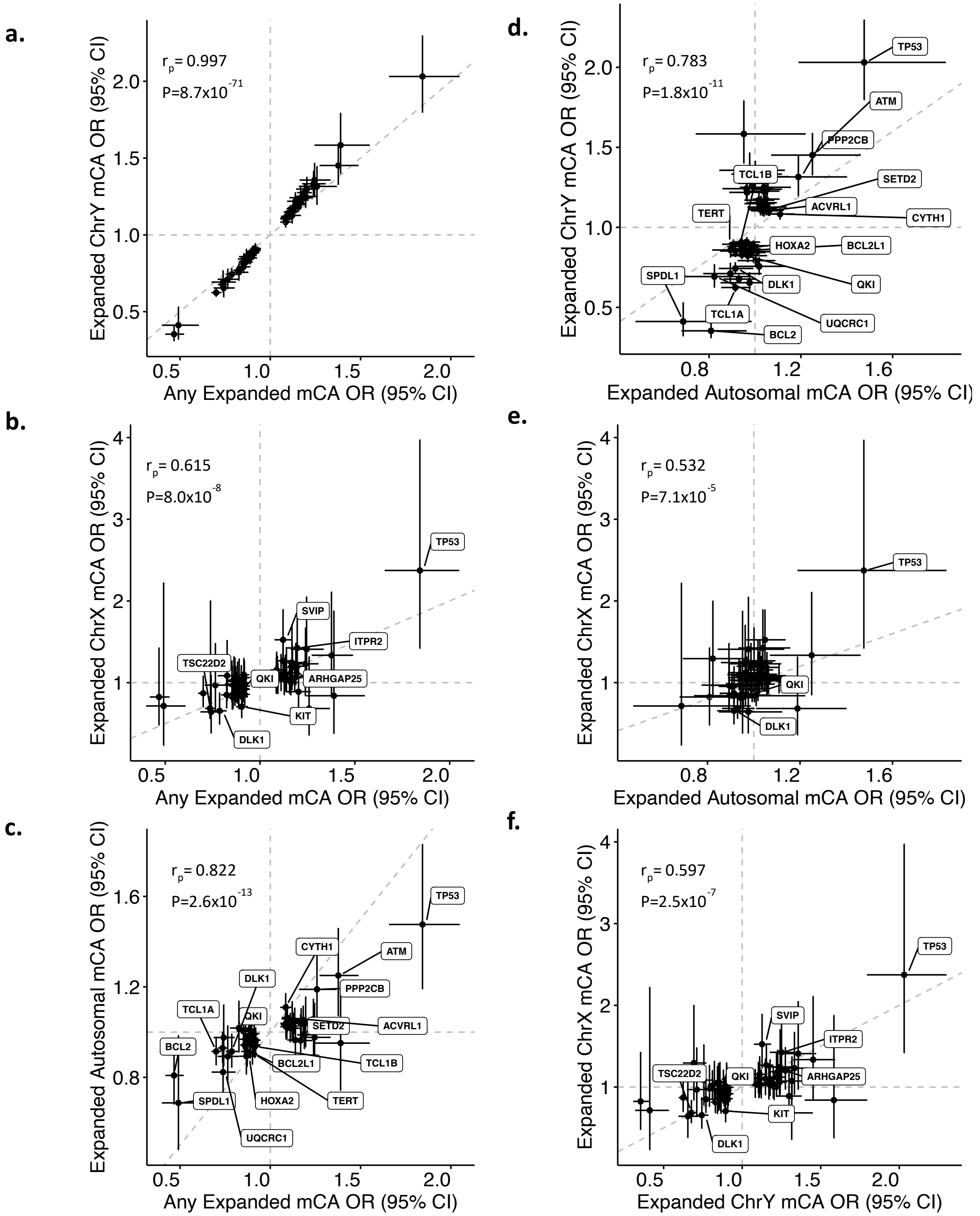


**Supplementary Note 16:** Correlated associations of 63 independent genome-wide significant variants associated with expanded mCAs (from Supplementary Data 3) between different mCA categories (expanded autosomal mCAs, expanded ChrX mCAs, expanded ChrY mCAs) in the UKB. Across all panels except for panel (a), the labeled genes represent genes attributed to variants that have P<0.05 across the mCA categories in both axes. mCA = mosaic chromosomal alterations, $r_p$ = Pearson correlation

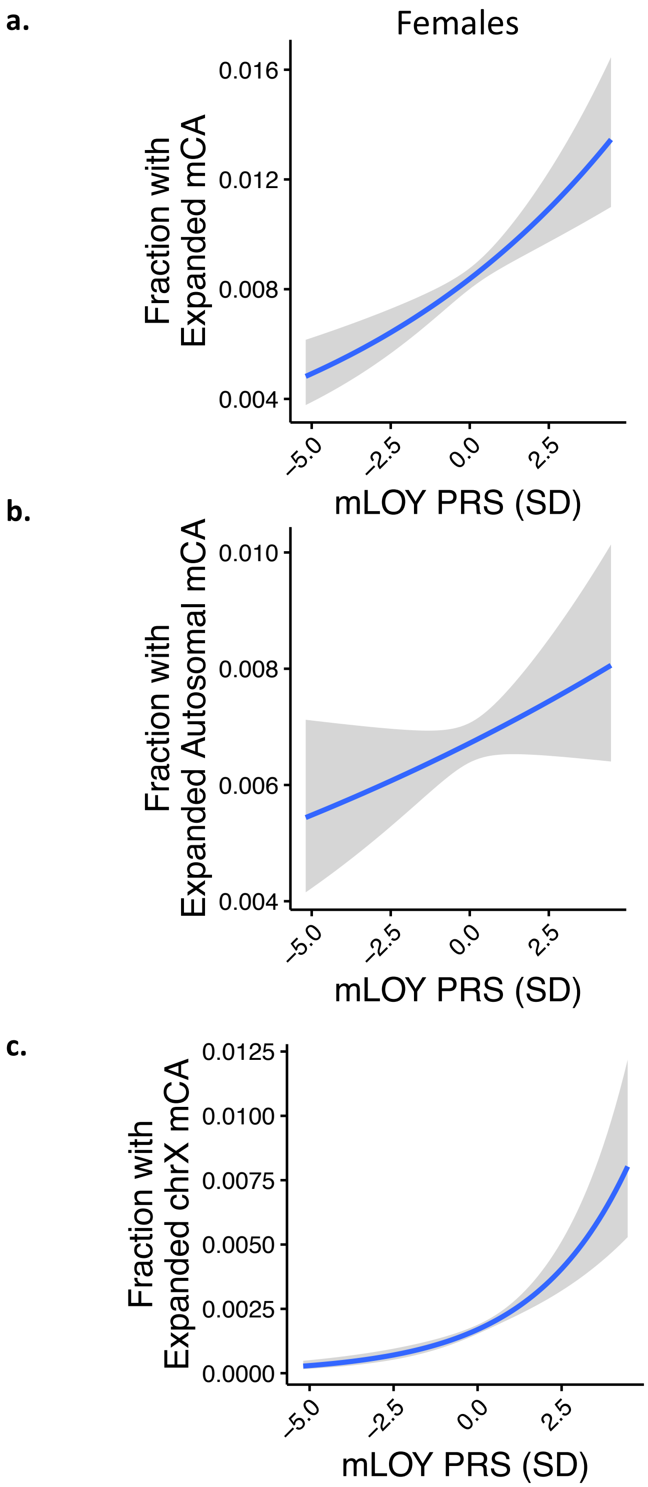


**Supplementary Note 17:** Association of a mLOY PRS consisting of 156 previously identified[1] independent genome-wide significant variants associated with mLOY, with different expanded mCA categories in UKB Females. mCA = mosaic chromosomal alterations, mLOY = mosaic Loss-of-chromosome Y, PRS = polygenic risk score.

| | ***UK Biobank*** | ***MGB Biobank*** | ***FinnGen**** | ***Biobank Japan*** |
|---|---|---|---|---|
| *N* | 444,199 | 22,461 | 175,690 | 125,541 |
| *Age of DNA collection (mean (SD))* | 56.5 (8) | 55.0 (16.8) | 53.4 (18.4) | 64.6 (12.4) |
| *Sex (Male (%))* | 204,579 (46.1%) | 10,306 (45.9%) | 71,000 (40.4) | 72,186 (57.5%) |
| *Prior or Current Smoker (%)* | 188,875 (45.0%) | 9,094 (40.5%) | 30,554 (42.7) | 66,913 (53.3%) |
| *Race* | White: 417,828 (94.1%)<br>Asian: 10,277 (2.3%)<br>Black: 7,173 (1.6%)<br>Mixed: 2,634 (0.6%)<br>Other: 4,160 (0.9%)<br>Unknown 187 (0.04%) | White: 18,933 (84.3%)<br>Asian: 569 (2.5%)<br>Black: 1,056 (4.7%)<br>Other: 744 (3.3%)<br>Unknown: 1,159 (5.2%) | White: 175,690 (100%) | Asian: 125,541 (100%) |
| *BMI (mean (SD))* | 27.4 (4.8) | 28.5 (6.2) | NA | 23.4 (3.7) |
| *Prevalent Solid Cancer* | 66,551 (15.0%) | 6,080 (27.1%) | 31,855 (18.1%) | 25,987 (20.7%) |
| *Prevalent Type 2 Diabetes Mellitus* | 10,835 (2.4%) | 1,782 (7.9%) | 22,326 (13.2%) | 31,636 (25.2%) |
| *Prevalent Coronary Artery Disease* | 25,287 (5.7%) | 3,908 (17.4%) | 19,474 (11.1%) | 23,099 (18.4%) |
| *Prevalent Hypertension* | 129,888 (29.2%) | 11,010 (49.0%) | NA | 37,913 (30.2%) |
| *Prevalent Hypercholesterolemia* | 66,483 (15.0%) | 9,881 (44.0%) | 8,583 (5.2%) | 35,026 (27.9%) |

**Supplementary Table 1:** Baseline summary statistics across the UK Biobank, MGB Biobank, FinnGen, and Biobank Japan among individuals analyzed.
* of note, 104,175 individuals from FinnGen had missing smoking status; BMI and hypertension were not available
BMI = body mass index; MGB = Mass General Brigham; SD = standard deviation.

| | *UK Biobank* | *MGB Biobank* | *FinnGen* | *Biobank Japan* |
|---|---|---|---|---|
| *N* | 444,199 | 22,461 | 175,690 | 125,541 |
| *Any mCA(%)* | 66,011 (14.9) | 3,784 (16.8) | 22,040 (12.5) | NA |
| *Autosomal mCA (%)* | 15,350 (3.5) | 1,025 (5.2) | 3,164 (2.0) | 20,440 (16.3) |
| *ChrX (%)* | 12,265 (5.1) | 820 (7.0) | 7,058 (6.8) | NA |
| *ChrY (%)* | 41,284 (20.1) | 2,201 (22.0) | 12,599 (18.0) | NA |
| *Any expanded mCA (%)* | 12,398 (3.2) | 1,026 (5.2) | 9,558 (5.9) | NA |
| *Expanded autosomal mCA (%)* | 2,985 (0.8) | 337 (1.8) | 1,620 (1.0) | 1,676 (1.3%) |
| *Expanded ChrX (%)* | 397 (0.2) | 44 (0.2) | 479 (0.5) | NA |
| *Expanded ChrY (%)* | 9168 (4.5) | 669 (3.4) | 7663 (11.8) | NA |

**Supplementary Table** 2**:** mCA counts by cohort.
Chr = chromosome; mCA = mosaic chromosomal alteration; MGB = Mass General Brigham; N = number.

| | *OR* | *Lower 95% CI* | *Upper 95% CI* | *P* |
|---|---|---|---|---|
| *Age* | 0.92 | 0.86 | 1.005 | 0.068 |
| *Age*$^2$ | 1.0013 | 1.0006 | 1.002 | 2.84E-05 |
| *Sex (Male)* | 1.44 | 1.33 | 1.56 | 1.33E-19 |
| *Prior or Current Smoking* | 1.09 | 1.01 | 1.18 | 0.03 |
| *Prevalent Solid Cancer* | 0.95 | 0.82 | 1.09 | 0.44 |
| *Prevalent Type 2 Diabetes Mellitus* | 1.05 | 0.84 | 1.32 | 0.66 |

**Supplementary Table 3:** Association of potential risk factors with expanded autosomal mCAs in the UK Biobank.
CI = confidence interval; OR = odds ratio

| *Phenotype* | *Exposure* | *Sex* | *HR* | *P* | *Controls (N)* | *Incident Cases (N)* | *Incident Cases with mCA (N)* | *Controls with mCA (N)* |
|---|---|---|---|---|---|---|---|---|
| *Nervous System Infection* | Autosomal mCA | Male | 3.71 | 0.0085 | 39,978 | 17 | 8 | 6,298 |
| *Meningitis or Encephalitis* | | Male | 3.71 | 0.0085 | 39,978 | 17 | 8 | 6,298 |
| *Pneumonia* | | All | 1.16 | 0.023 | 72,317 | 1,299 | 334 | 10,485 |
| *Nervous System Infection* | | All | 2.81 | 0.025 | 72,317 | 21 | 8 | 10,485 |
| *Meningitis or Encephalitis* | | All | 2.81 | 0.025 | 72,317 | 21 | 8 | 10,485 |
| *Respiratory System Infection* | | All | 1.15 | 0.028 | 72,317 | 1,353 | 346 | 10,485 |
| *Any Infection* | | All | 1.12 | 0.037 | 72,317 | 1,998 | 476 | 10,485 |
| *Pneumonia* | | Male | 1.17 | 0.039 | 39,978 | 928 | 259 | 6,298 |
| *Endocarditis or Myocarditis* | | Female | 2.46 | 0.043 | 32,339 | 25 | 8 | 4,187 |
| *Respiratory System Infection* | | Male | 1.16 | 0.050 | 39,978 | 971 | 269 | 6,298 |
| *Sepsis* | Expanded | All | 2.04 | 0.050 | 72,317 | 276 | 8 | 753 |
| *Sepsis* | Autosomal mCA | All | 2.04 | 0.050 | 72,317 | 276 | 8 | 753 |

**Supplementary Table 4:** Association of mCAs with mortality from incident infection in Biobank Japan. Suggestive associations (P<0.05) are presented among individuals without antecedent cancer prior to the infection phenotype.
HR = hazard ratio; mCA = mosaic chromosomal alteration; N = number

| *Incident Sepsis Multivariate Model Component* | *HR* | *P* | *Lower 95% CI* | *Upper 95% CI* |
|---|---|---|---|---|
| *Expanded Autosomal mCA* | 2.00 | 2.64E-08 | 1.56 | 2.54 |
| *Antecedent Cancer Prior to Incident Sepsis* | 5.49 | <1E-300 | 5.09 | 5.93 |
| *Age* | 1.02 | 0.64 | 0.95 | 1.09 |
| *Age*$^2$ | 1.00 | 0.41 | 1.00 | 1.00 |
| *Sex (Male)* | 1.27 | 9.56E-11 | 1.18 | 1.37 |
| *current cigar pipe smoker, former cigarette smoker* | 1.90 | 0.011 | 1.16 | 3.12 |
| *current cigar pipe smoker, not former cigarette smoker* | 1.81 | 0.077 | 0.94 | 3.49 |
| *current cigarette smoker, 10 to <20/day* | 1.83 | 1.79E-10 | 1.52 | 2.20 |
| *current cigarette smoker, 20 to <40/day* | 2.06 | 8.23E-14 | 1.70 | 2.49 |
| *current cigarette smoker, <10/day* | 1.39 | 0.047 | 1.00 | 1.92 |
| *current cigarette smoker, ≥40/day* | 3.33 | 3.65E-05 | 1.88 | 5.89 |
| *current occasional smoker, smoked cigarettes daily in past, <20/day* | 1.37 | 0.19 | 0.86 | 2.18 |
| *current occasional smoker, smoked cigarettes daily in past, ≥20/day* | 1.83 | 0.01 | 1.13 | 2.96 |
| *current occasional smoker, smoked cigars or pipes daily in past* | 0.51 | 0.50 | 0.07 | 3.60 |
| *current occasional smoker, smoked ≥100 cigarettes in lifetime* | 0.99 | 0.94 | 0.70 | 1.39 |
| *former cigarette smoker, <20/day, quit 1-5 year ago* | 1.14 | 0.48 | 0.80 | 1.63 |
| *former cigarette smoker, <20/day, quit 10-20 year ago* | 1.15 | 0.27 | 0.90 | 1.46 |
| *former cigarette smoker, <20/day, quit 5-10 year ago* | 1.29 | 0.11 | 0.94 | 1.78 |
| *former cigarette smoker, <20/day, quit <1 year ago* | 1.90 | 0.06 | 0.98 | 3.65 |
| *former cigarette smoker, <20/day, quit ≥20 year ago* | 1.08 | 0.34 | 0.92 | 1.26 |
| *former cigarette smoker, ≥20/day, quit 1-5 year ago* | 2.25 | 1.07E-13 | 1.82 | 2.79 |
| *former cigarette smoker, ≥20/day, quit 10-20 year ago* | 1.54 | 7.00E-07 | 1.30 | 1.82 |
| *former cigarette smoker, ≥20/day, quit 5-10 year ago* | 1.64 | 1.11E-05 | 1.31 | 2.04 |
| *former cigarette smoker, ≥20/day, quit <1 year ago* | 2.08 | 0.0064 | 1.23 | 3.53 |
| *former cigarette smoker, ≥20/day, quit ≥20 year ago* | 1.25 | 0.00080 | 1.10 | 1.42 |
| *former daily cigar pipe smoker* | 1.23 | 0.19 | 0.91 | 1.66 |
| *former occasional cigarette smoker, lifetime cigarette smoking unknown* | 1.07 | 0.76 | 0.70 | 1.63 |
| *former occasional cigarette smoker, smoked <100 cigarettes in lifetime* | 1.27 | 0.085 | 0.97 | 1.66 |
| *former occasional cigarette smoker, smoked ≥100 cigarettes in lifetime* | 1.02 | 0.72 | 0.90 | 1.17 |
| *Missing smoking* | 1.55 | 0.0085 | 1.12 | 2.15 |
| *BMI (SD)* | 1.21 | 7.08E-31 | 1.17 | 1.25 |
| *Prevalent Type 2 Diabetes Mellitus* | 1.84 | 1.75E-16 | 1.59 | 2.12 |
| *Leukocyte count (SD)* | 1.31 | 1.38E-07 | 1.19 | 1.45 |
| *Lymphocyte count (SD)* | 0.71 | 4.57E-07 | 0.63 | 0.81 |
| *Lymphocyte percentage (SD)* | 1.12 | 0.054 | 1.00 | 1.27 |
| *PC1* | 0.98 | 0.18 | 0.96 | 1.01 |
| *PC2* | 0.99 | 0.42 | 0.97 | 1.01 |
| *PC3* | 0.99 | 0.59 | 0.97 | 1.02 |
| *PC4* | 0.99 | 0.37 | 0.98 | 1.01 |
| *PC5* | 1.01 | 0.07 | 1.00 | 1.01 |
| *PC6* | 1.01 | 0.46 | 0.99 | 1.03 |
| *PC7* | 0.99 | 0.23 | 0.97 | 1.01 |
| *PC8* | 1.01 | 0.58 | 0.99 | 1.03 |
| *PC9* | 1.00 | 0.58 | 0.99 | 1.01 |
| *PC10* | 1.00 | 0.61 | 0.99 | 1.02 |

**Supplementary Table 5:** Sensitivity analysis of incident sepsis association in the UK Biobank with the addition of a 25-factor smoking covariate, leukocyte count, lymphocyte count and percentage, BMI, and prevalent type 2 diabetes mellitus.
BMI = body mass index; CI = confidence interval; HR = hazard ratio; PC = principal component; SD = standard deviation

| *Outcome* | *Population of people with cancer prior to infection* | *HR* | *P* | *Lower 95% CI* | *Upper 95% CI* | *Cases (N)* | *Controls (N)* | *Cases with mCA (N)* | *Controls with mCA (N)* |
|---|---|---|---|---|---|---|---|---|---|
| *Sepsis* | Prevalent Solid Cancer | 1.86 | 0.0097 | 1.16 | 2.96 | 1258 | 64921 | 20 | 521 |
| | Incident Solid Cancer Prior to Infection | 0.76 | 0.44 | 0.38 | 1.53 | 1619 | 51867 | 10 | 361 |
| | Incident Hematologic Cancer Prior to Infection | 0.98 | 0.88 | 0.77 | 1.25 | 833 | 2864 | 83 | 312 |
| | Incident Hematologic Cancer and Prevalent Solid Cancer Prior to Infection | 0.94 | 0.85 | 0.52 | 1.72 | 144 | 546 | 15 | 63 |
| | **Any Cancer Prior to Infection** | **2.82** | **5.28E-22** | **2.28** | **3.48** | **3575** | **119106** | **99** | **1131** |
| *Pneumonia* | Prevalent Solid Cancer | 1.68 | 0.0057 | 1.16 | 2.43 | 2382 | 62325 | 40 | 480 |
| | Incident Solid Cancer Prior to Infection | 1.33 | 0.18 | 0.87 | 2.03 | 2369 | 49466 | 24 | 323 |
| | Incident Hematologic Cancer Prior to Infection | 1.19 | 0.18 | 0.92 | 1.54 | 655 | 2886 | 80 | 300 |
| | Incident Hematologic Cancer and Prevalent Solid Cancer Prior to Infection | 1.73 | 0.076 | 0.94 | 3.18 | 119 | 528 | 16 | 55 |
| | **Any Cancer Prior to Infection** | **2.26** | **5.08E-17** | **1.86** | **2.73** | **5295** | **114149** | **130** | **1048** |

**Supplementary Table 6:** Sensitivity analysis of incident sepsis and pneumonia association in the UK Biobank among populations of individuals with different types of cancer prior to incident infection, where solid cancer is defined as any non-hematologic cancer. Other covariates in the model included age, age$^2$, sex, smoking status, and PC1-10 of ancestry.
CI = confidence interval; HR = hazard ratio; mCA = mosaic chromosomal alteration; N = number
